# Mpox severity and mortality in the DRC: a systematic review and meta-analysis (1970-2024)

**DOI:** 10.1101/2025.04.07.25325410

**Authors:** Fabrice Zobel Lekeumo Cheuyem, Armel Zemsi, Jonathan Hangi Ndungo, Chabeja Achangwa, Davy Roméo Takpando-le-grand, Jessy Goupeyou-Youmsi, Solange Dabou, Mohamadou Adama, Ariane Nouko, Marie Angèle Mbang, Edwige Omona Guissana, Rick Tchamani, Vanessa Monique Mballa Assiene, Ethel Ambo Eno, Constantine Tanywe Asahngwa, Saralees Nadarajah

## Abstract

**Background:** Mpox, caused by the Mpox virus, is a zoonotic disease historically endemic in Central and West Africa. The Democratic Republic of Congo (DRC) bears the highest burden, with evolving epidemiology and significant public health challenges. Understanding severity and mortality trends is critical for global control efforts.

**Methods:** This systematic review and meta-analysis followed PRISMA guidelines to synthesize evidence on Mpox severity and mortality in the DRC from 1970 to 2024. We searched PubMed, ScienceDirect, Scopus, Web of Science identifying 19 eligible studies after screening 5443 records. Data on severity and death among suspected and confirmed cases were extracted. Random-effects models addressed high heterogeneity, and subgroup analyses examined temporal trends, regional differences, and healthcare settings. Meta-regression explored sources of heterogeneity, adjusting for study year, region, and setting.

**Results:** We included a total of 19 study reports. The pooled severity rate among 3,280 confirmed cases was 39.47% (95% confidence interval [CI]: 29.11-50.86), declining from 46.34% (95% CI: 37.07-55.87) pre-2022 to 25.83% (95% CI: 11.16-49.11) in 2022-2024. Northern (43.82%; 95% CI: 37.81-50.03) and Central (63.89%; 95% CI: 57.10-70.30) regions had higher severity than Eastern regions (26.04%; 95% CI: 8.77-56.31). Case fatality rates (CFRs) were 1.70% (95% CI: 0.72-3.95) for suspected cases and 2.46% (95% CI: 0.65-8.85) for confirmed cases, with Western DRC disproportionately affected (22.03%; 95% CI: 13.25-34.34). Community settings showed higher CFRs (3.69%; 95% CI: 0.78-15.81) than hospitals (0.77%; 95% CI: 0.32-1.84), underscoring healthcare access disparities. Meta-regression confirmed study year (p = 0.044) as significant predictors of outcomes case severity heterogeneity and study year (*p* = 0.016), region (*p* = 0.003) and setting (*p* = 0.041), with mortality declining over time but remaining elevated in resource-limited areas.

**Conclusion:** Mpox continues to impose a substantial burden in the DRC, with high severity and mortality rates, particularly in Western/Central regions and community settings. The observed temporal decline in severity and CFRs suggests the impact of strengthened surveillance and healthcare capacity. However, regional disparities persist might be due to inequitable healthcare access and potential differences in viral clades. Targeted interventions, including vaccination in high-risk areas, community education, and healthcare system strengthening are urgently needed.

## Background

Monkeypox (Mpox), caused by the Mpox virus, is a zoonotic infection that has garnered increasing global attention due to its evolving epidemiology [1]. The Mpox virus belongs to the *Orthopoxvirus* genus, which also includes variola virus (the causative agent of smallpox), vaccinia virus (used in the smallpox vaccine), and cowpox virus [2]. Mpox virus is a double-stranded DNA virus with two distinct genetic clades: clade 1 (formerly known as the Congo Basin clade) and clade 2 (formerly the West African clade). Clade 1 has historically been associated with more severe disease and higher mortality rates compared to Clade 2 [3].

While historically concentrated in Central and West Africa, recent outbreaks have demonstrated its capacity to spread beyond endemic regions, posing a significant threat to global public health [4]. In the Democratic Republic of Congo (DRC), Mpox has been recognized as a public health threat since 1997, leading to the establishment of the National Program for the Control of Mpox and Hemorrhagic Fevers and the designation of Mpox as an epidemic-prone disease requiring immediate and mandatory notification [5]. The retrospective analysis of national routine surveillance data from 2010 to 2023 shows a concerning rise in Mpox incidence and geographical spread in the DRC. Over that period, the incidence rate rose from 2.97 to 11.46 per 100,000 people, with 60,967 suspected cases and 1,798 deaths reported, and children under 5 years were particularly affected [6].

The clinical pattern of Mpox infection is a characteristic skin rash, progressing through stages of macules, papules, vesicles, and pustules, eventually forming crusts [7]. This rash can be accompanied by systemic symptoms such as fever, headache, lymphadenopathy, and myalgia [8]. The severity of Mpox infection can vary widely, ranging from mild, self-limiting illness to severe or grave disease requiring hospitalization and leading to complications [9].

The transmission of Mpox virus to humans can occur through various routes [10]. Zoonotic transmission involves contact with infected animals, such as rodents and non-human primates [11,12]. Human-to-human transmission primarily occurs through close contact with infectious skin lesions, respiratory droplets, or contaminated materials. Sexual transmission has also been documented, particularly in recent outbreaks [3,13].

The epidemiology of Mpox has been significantly shaped by the global eradication of smallpox [1]. Smallpox vaccination, using vaccinia virus, provided cross-protective immunity against other *Orthopoxviruses*, including Mpox virus [14]. Following the cessation of smallpox vaccination campaigns, susceptibility to Mpox increased in the population, leading to a rise in Mpox incidence and a shift in its epidemiology [15].

Socioeconomic factors play a crucial role in shaping Mpox epidemiology in the DRC [8]. Poverty, limited access to healthcare, inadequate sanitation, and cultural practices can increase the risk of Mpox transmission and exacerbate its impact [16]. In addition, social instability and conflict can disrupt healthcare services and hinder disease control efforts [17].

The healthcare system in the DRC faces significant challenges, including limited infrastructure, shortages of healthcare workers, inadequate diagnostic capacity, and logistical difficulties in reaching remote areas [16]. These challenges can result in delayed diagnosis, inadequate treatment, and increased mortality from Mpox [18].

In light of the ongoing public health importance of Mpox in the DRC and its potential for global spread, it is imperative to enhance our understanding of the disease’s severity and mortality patterns. Accurate and up-to-date information on these key epidemiological parameters is essential for informing evidence-based public health interventions and allocating resources effectively. This systematic review and meta-analysis aimed to address this critical need by synthesizing available data on Mpox severity and case fatality rates (CFRs) in the DRC. By examining trends over time and variations across different settings and populations, this study provides valuable insights into the current and historical burden of Mpox in the region, contributing to global efforts to control and mitigate the impact of this emerging infectious disease.

## Methods

### Study design

This study was conducted to assess the epidemiology of the Mpox outbreak in the DRC from 1970 to 2024, as this was the first meta-analysis assessing disease severity and mortality in this most endemic focus. The study results are reported based on the Preferred Reporting Items for Systematic Review and Meta-analysis (PRISMA) guidelines [19].

### Study setting

The DRC is the largest country in Sub-Saharan Africa, occupying a vast land area of approximately 2,345,409 km^²^ [20]. Administratively, the DRC is divided into 26 provinces, including Bas-Uélé, Équateur, Haut-Katanga, Haut-Lomami, Haut-Uélé, Ituri, Kasaï, Kasaï-Central, Kasaï-Oriental, Kinshasa, Kongo Central, Kwango, Kwilu, Lomami, Lualaba, Mai-Ndombe, Maniema, Mongala, Nord-Kivu, Nord-Ubangi, Sankuru, Sud-Kivu, Sud-Ubangi, Tanganyika, Tshopo, and Tshuapa. The country’s health system is organized into central, intermediate, and operational levels. At the operational level, there are 519 health zones, 393 general referral hospitals, and 8,504 health areas [21]. The population is estimated to be over 105 million, with a median age of 16.7 years, a population growth rate of 3.1%, and a life expectancy of 61.6 years [22]. The health landscape is plagued by recurrent disease outbreaks, such as mpox, yellow fever, polio, cholera, measles, and Ebola [23]. The majority of the population is Christian, with over 93% identifying as such, of which 42% are Catholic [24]. The majority of the population is Christian, with over 93% identifying as such, of which 42% are Catholics [25]. The DRC has a long history of armed groups steering civil unrest, resulting in a complex humanitarian emergency [26].

### Eligibility criteria

#### Inclusion criteria

This systematic review and meta-analysis included community- and hospital-based studies (cross-sectional, surveillance reports, cohort) that provided data on Mpox severity and/or mortality rates in the Democratic Republic of Congo (DRC). Studies were eligible if they reported clinical outcomes (e.g., severe cases, deaths) or epidemiological surveillance data (e.g., case fatality rates). No restrictions were applied on publication year; however, only articles published in English or French were considered.

#### Exclusion criteria

Studies were excluded if they were single case reporting, reviews, or commentaries without original data, or if they focused on other viruses or unrelated outcomes (e.g., genetic analyses without clinical correlates). Duplicate publications were removed following title and abstract screening. Additionally, studies lacking clearly defined severity or mortality measures were excluded.

### Article searching strategy

A methodical investigation of digital repositories, encompassing PubMed, Web of Science, Scopus, Embase, CINAHL and ScienceDirect, was performed to locate published research. The search approach involved examining the words within the title and abstract of each study. A synthesis of keywords and Medical Subject Headings (MeSH) vocabulary was utilized, employing Boolean logic operators (“AND” and “OR”) to narrow the search for each database used (Additional File 1, Supplementary Table 2). To ensure thoroughness, additional search was conducted on Google Scholar to identify additional relevant publications not found in electronic databases. Furthermore, the reference lists of identified studies were screened to identify further relevant articles. The concluding search was performed on 29 June, 2025.

### Data extraction

Data were extracted from all eligible articles using a predefined Microsoft Excel 2016 form to collect study characteristics. The data extraction checklist included the first author’s name, study year, region, setting, number of suspected, confirmed, severe and death Mpox cases. Two authors independently assessed each article for relevance and quality (F.Z.L.C., A.Z.). Discrepancies between reviewers were resolved through discussion with a third reviewer to achieve consensus (S.N).

### Data quality assessment

The quality of included studies was assessed using the Joanna Briggs Institute (JBI) quality assessment tool [27]. Risk of bias was evaluated using predefined criteria based on the study design. For cross-sectional studies, assessment criteria encompassed studies clear definition of inclusion criteria, comprehensive descriptions of study subjects and settings, validity and reliability of exposure measurements, use of objective, standardized criteria for outcome assessment, identification potential confounding factors, implementation of appropriate strategies to address them, and the appropriateness of statistical methods. For case reports, criteria included: clear description of patient demographic characteristics, patient history and timeline, current clinical condition on presentation, diagnostic tests/assessment methods and results, interventions/treatment procedures, post-intervention clinical condition, adverse/unanticipated events, and takeaway lessons. Each criterion was scored as 1 (yes) or 0 (no or unclear). The overall risk of bias was categorized as low (>50%), moderate (>25-50%), or high (≤25%).

### Outcome measurement

The primary outcomes of this systematic review and meta-analysis were Mpox severity and CFR. Mpox severity rate was determined by calculating the proportion of participants exhibiting severe or grave clinical manifestations of Mpox out of the total number of confirmed cases. CFR was calculated by dividing the number of deaths observed within the population of suspected or confirmed cases.

### Operational definition

Severe or grave cases (requiring hospitalization), were defined by the presence of one or more of the following: extensive lesions (more than 100 lesions), hemorrhagic or pustular lesions, mucosal involvement (oral, genital, conjunctival), or systemic complications, including high fever (>39°C), altered general state (bedridden), sepsis, encephalitis, secondary bacterial infections, hypotension, septic shock, or severe dehydration, and keratitis leading to potential corneal scarring and blindness [8,28]. A suspected monkeypox case was defined as an individual presenting with a vesicular or pustular rash characterized by deep-seated, firm pustules, and at least one of the following symptoms: fever preceding the eruption, lymphadenopathy (inguinal, axillary, or cervical), or pustules or crusts on the palms of the hands or soles of the feet. A case was defined as laboratory-confirmed Mpox if at least one specimen yielded a positive result in the *Orthopoxvirus*-specific assay, or Mpox-specific real-time PCR, or Mpox in culture [8].

### Statistical analysis and synthesis

Heterogeneity between studies was assessed using the I^²^ statistic and categorized as low (<25%), moderate (25-75%), or high (>75%). Subgroup analyses were conducted based on study period, region, setting, and disease burden. A random-effects model was employed when heterogeneity exceeded 50%. Furthermore, meta-regression was performed to investigate whether study characteristics explained the variability in results across studies. The timeframes for the study period subgroup analysis were specifically defined as before and after the 2022 global outbreak.

Only study variables with meaningful and practical categories were included in the analyses. Univariable and multivariable meta-regression models were used to assess whether severity and CFRs varied according to the selected explanatory variable categories. Generalized Linear Mixed Models (GLMM), coupled with the Probit-Logit Transformation (PLOGIT) were used as they are ideal for meta-analyses of binary or proportion data as they directly model binomial outcomes, effectively account for between-study heterogeneity using random effects, and inherently handle studies with 0% or 100% events without requiring continuity corrections [33]. Statistical significance was set at 5%. All analyses were performed using the ‘meta’ package in R Statistics version 4.4.2 [34].

### Publication bias and sensitivity test

Publication bias was assessed visually using a funnel plot. Symmetry of the inverted funnel shape suggested the absence of publication bias. To further investigate potential bias, Egger’s regression and Begg’s rank correlation tests were performed, with statistical significance set at 5%. Sensitivity analysis was conducted by iteratively excluding one study at a time to evaluate the robustness of the finding.

## Results

A total of 5443 records were retrieved from the database search (*n* = 5438) and from other sources (*n* = 5). After removing 732 duplicate records, 4706 records remained. Titles/abstracts, followed by full text articles were then screened for eligibility. Ultimately, 19 study reports met the eligibility criteria and were included in the meta-analysis (Fig. 1).

**Fig. 1.**
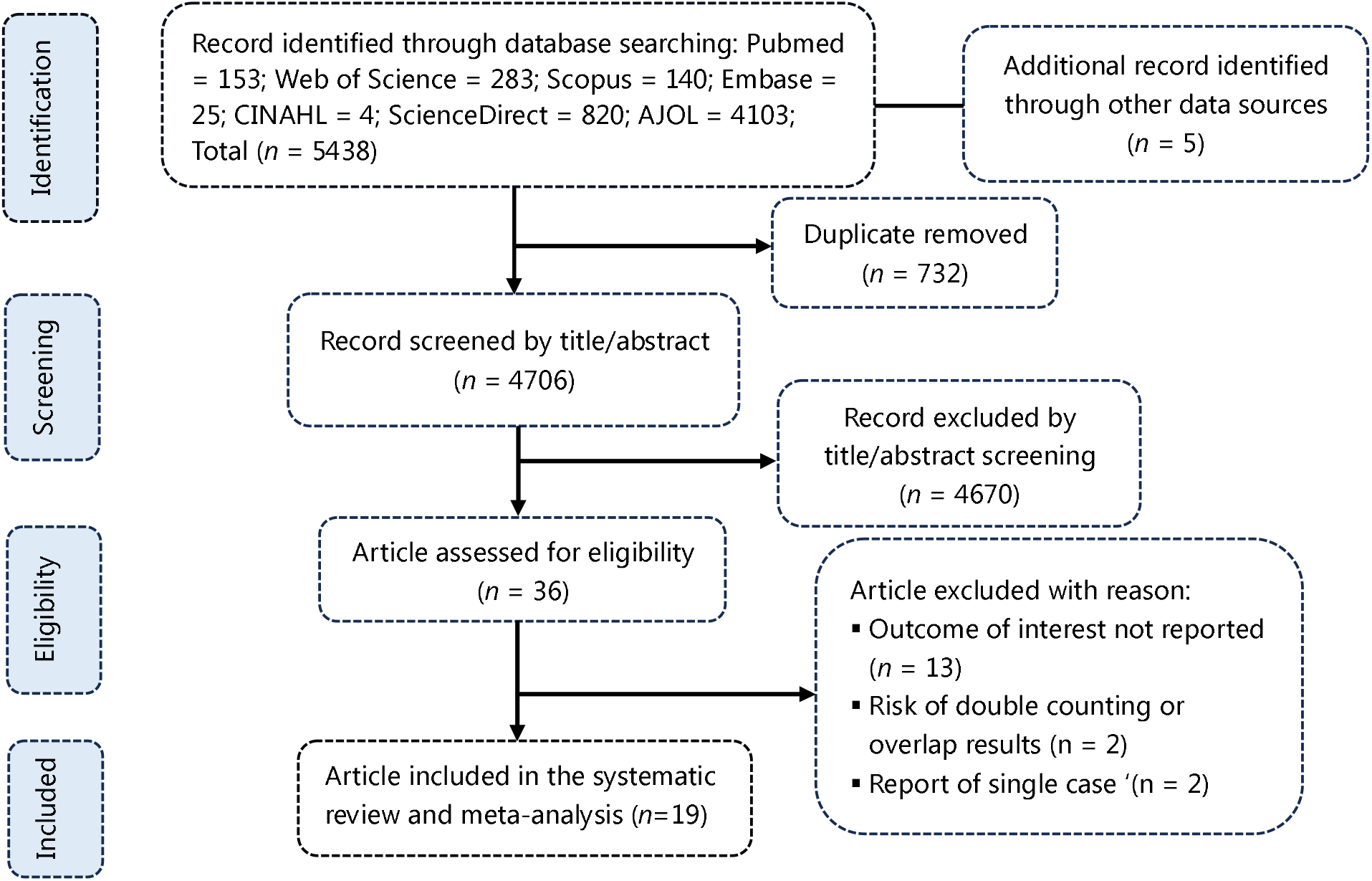
PRISMA diagram flow from study identification to inclusion in the meta-analysis

### Studies selection

### Characteristic of reports included

A comprehensive analysis included 22 studies. Those studies were conducted between 1970 and 2024 and involved population from community, hospital settings or both across various regions in DRC. Most of studies were surveillance reports with cross-sectional design for data collection and description (*n* = 16) (Additional Files 1, Supplementary Table 1)

### Disease severity among confirmed Mpox cases

The overall pooled Mpox severity rate was 39.47% (95% confidence interval [CI]: 29.11-50.86; *I*^²^ = 93.9%, *p* < 0.001) (Fig. 2). Subgroup analysis by study period underscores a decline in the Mpox severity rate from 46.34% (95% CI: 37.07-55.87; n = 8 studies) before 2022 to 11.16% (95% CI: 11.16-49.11; n = 5 studies) from 2022-2024. The highest severity rate was observed in the Central region of Sankuru (63.89%; 95% CI: 57.10-70.30; n = 1 study), while the lowest was in the Eastern regions of South and North Kivu (26.04%; 95% CI: 8.77-56.31; n = 3 studies); a significant difference in severity rates was noted across regions (*p* < 0.001). The severity rate was higher in studies conducted within hospital settings (43.21%; 95% CI: 19.15-70.96; n = 2 studies) compared to community settings (38.68%; 95% CI: 27.74-50.90; n = 11 studies). Sites with the highest burden of confirmed Mpox cases (≥ 200) exhibited the highest severity rate (40.52%; 95% CI: 31.62-50.10; n = 7 studies) (Table 1 and Additional Files 2, Supplementary Fig. 1-4).

**Table 1.** Subgroup meta-analysis of Mpox severity rate pooled estimates in DRC, 1970-2024.

| Subgroup | Confirmed cases | Event rate<sup>1</sup> (%) | 95% CI limits<sup>1</sup> |  | Number of studies | Heterogeneity statistic<sup>1</sup> |  |
| --- | --- | --- | --- | --- | --- | --- | --- |
|  |  |  | Lower | Upper |  | I<sup>2</sup>(%) | p-value |
| Study period<sup>2</sup> |  |  |  |  |  |  |  |
| <2022 | 2497 | 46.34 | 37.07 | 55.87 | 8 | 93.2 | < 0.001 |
| 2022-2024 | 783 | 25.83 | 11.16 | 49.11 | 5 | 92.1 | < 0.001 |
| Region<sup>3</sup> |  |  |  |  |  |  |  |
| Northern | 251 | 43.82 | 37.81 | 50.03 | 2 | 0 | 0.581 |
| Eastern | 548 | 26.04 | 8.77 | 56.31 | 3 | 89.2 | < 0.001 |
| Western | 1983 | 42.71 | 31.17 | 55.10 | 6 | 92.6 | < 0.001 |
| Central | 216 | 63.89 | 57.10 | 70.30 | 1 | - | - |
| Nationwide | 282 | 39.01 | 33.28 | 44.97 | 1 | - | - |
| Setting |  |  |  |  |  |  |  |
| Community | 2633 | 38.68 | 27.74 | 50.90 | 11 | 90.7 | < 0.001 |
| Hospital | 647 | 43.21 | 19.15 | 70.96 | 2 | 98.8 | < 0.001 |
| Disease burden (confirmed cases) |  |  |  |  |  |  |  |
| < 200 | 197 | 36.81 | 16.13 | 63.82 | 6 | 88.5 | < 0.001 |
| ≥ 200 | 3083 | 40.52 | 31.62 | 50.10 | 7 | 96.1 | < 0.001 |

<sup>1</sup>Random effects model; CI: Confidence Interval; <sup>2</sup>Period before and after the global outbreak; <sup>3</sup>Eastern: North and South Kivu; Northern: Bas-ulé and Tshopo; Western/Central: Equateur Kwango, Sankuru, Tshuapa)

**Fig. 2.**
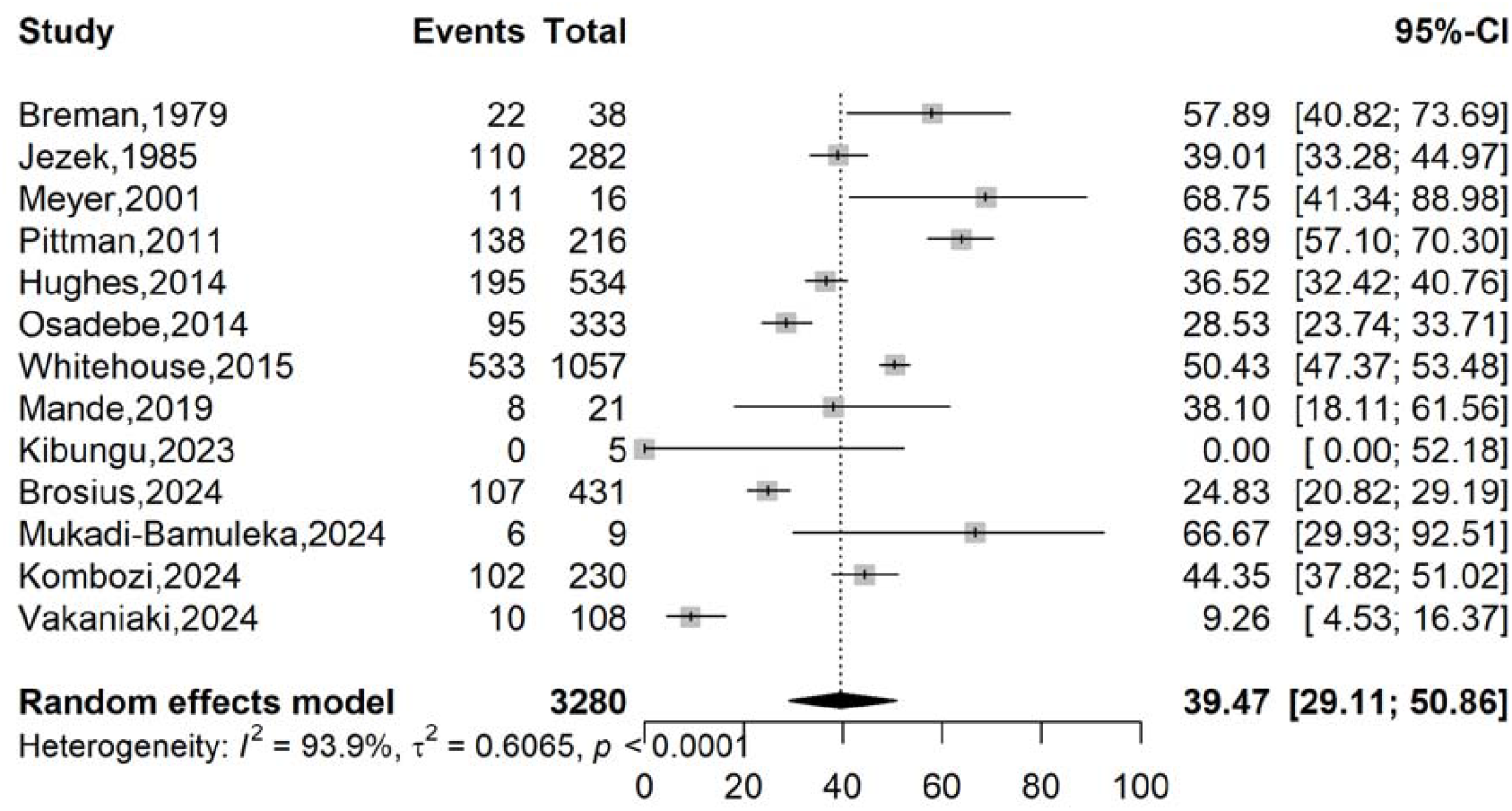
Pooled severity rate of Mpox case in DRC, 1970-2024

### Mortality among suspected Mpox cases

The pooled Case Fatality Rate (CFR) among suspected Mpox cases was 1.70% (95% CI: 0.70-3.95; *I*^²^ = 86.7%, *p* < 0.001) (Fig. 3). Subgroup analysis revealed a significant decreasing trend (p < 0.001) in CFR from 3.50% (95% CI: 1.60-7.51; n = 8 studies) before 2022 to 1.28% (95% CI: 0.66-1.90; n = 13 studies) from 2022-2024. The highest burden of mortality was recorded in Western regions (18.60%; 95% CI: 9.59-33.01; n = 2 studies), while the lowest was in Eastern regions (0.53%; 95% CI: 0.20-1.39%); this difference was statistically significant (*p* < 0.001). The CFR was higher within community settings (2.15%; 95% CI: 0.82-5.48%) compared to hospital settings (0.68%; 95% CI: 0.28-1.62%). Localities with the lowest burden of suspected cases (< 200) presented with a higher CFR (4.74%; 95% CI: 1.51-13.89%) (Table 2 and Additional Files 3, Supplementary Fig. 1-4).

**Table 2.** Subgroup meta-analysis of case fatality rate pooled estimates for suspected Mpox cases in DRC, 1970-2024.

| Subgroup | Suspected cases | Event rate<sup>1</sup> (%) | 95% CI limits<sup>1</sup> |  | Number of studies | Heterogeneity statistic<sup>1</sup> |  |
| --- | --- | --- | --- | --- | --- | --- | --- |
|  |  |  | Lower | Upper |  | I<sup>2</sup>(%) | p-value |
| Study period<sup>2</sup> |  |  |  |  |  |  |  |
| <2022 | 1420 | 3.50 | 1.60 | 7.51 | 8 | 85.6 | < 0.001 |
| 2022-2024 | 1732 | 0.35 | 0.16 | 0.77 | 5 | 0.0 | 0.747 |
| Region<sup>3</sup> |  |  |  |  |  |  |  |
| Western | 43 | 18.60 | 9.59 | 33.01 | 2 | 0.0 | 0.999 |
| Eastern | 761 | 0.53 | 0.20 | 1.39 | 3 | 94.5 | 0.753 |
| Nationwide | 1524 | 1.69 | 0.26 | 10.02 | 3 | 94.5 | < 0.001 |
| Central | 821 | 1.71 | 1.01 | 2.86 | 4 | 11.5 | 0.335 |
| Northern | 3 | 0.00 | 0.00 | 70.76 | 1 | - | - |
| Setting |  |  |  |  |  |  |  |
| Community | 2416 | 2.15 | 0.82 | 5.48 | 10 | 87.5 | < 0.001 |
| Hospital | 736 | 0.68 | 0.28 | 1.62 | 3 | 0.0 | 0.395 |
| Disease burden (suspected cases) |  |  |  |  |  |  |  |
| < 200 | 235 | 4.74 | 1.51 | 13.89 | 6 | 60.5 | 0.027 |
| ≥ 200 | 2917 | 1.13 | 0.43 | 2.89 | 7 | 91.3 | < 0.001 |

<sup>1</sup>Random effects model; CI: Confidence Interval; <sup>2</sup>Period before and after the global outbreak; <sup>3</sup>Eastern: North Kivu and South Kivu; Central: Kasai Oriental and Sankuru; Northern: North Ubangui; Western: Equateur and Kwango

**Fig. 3.**
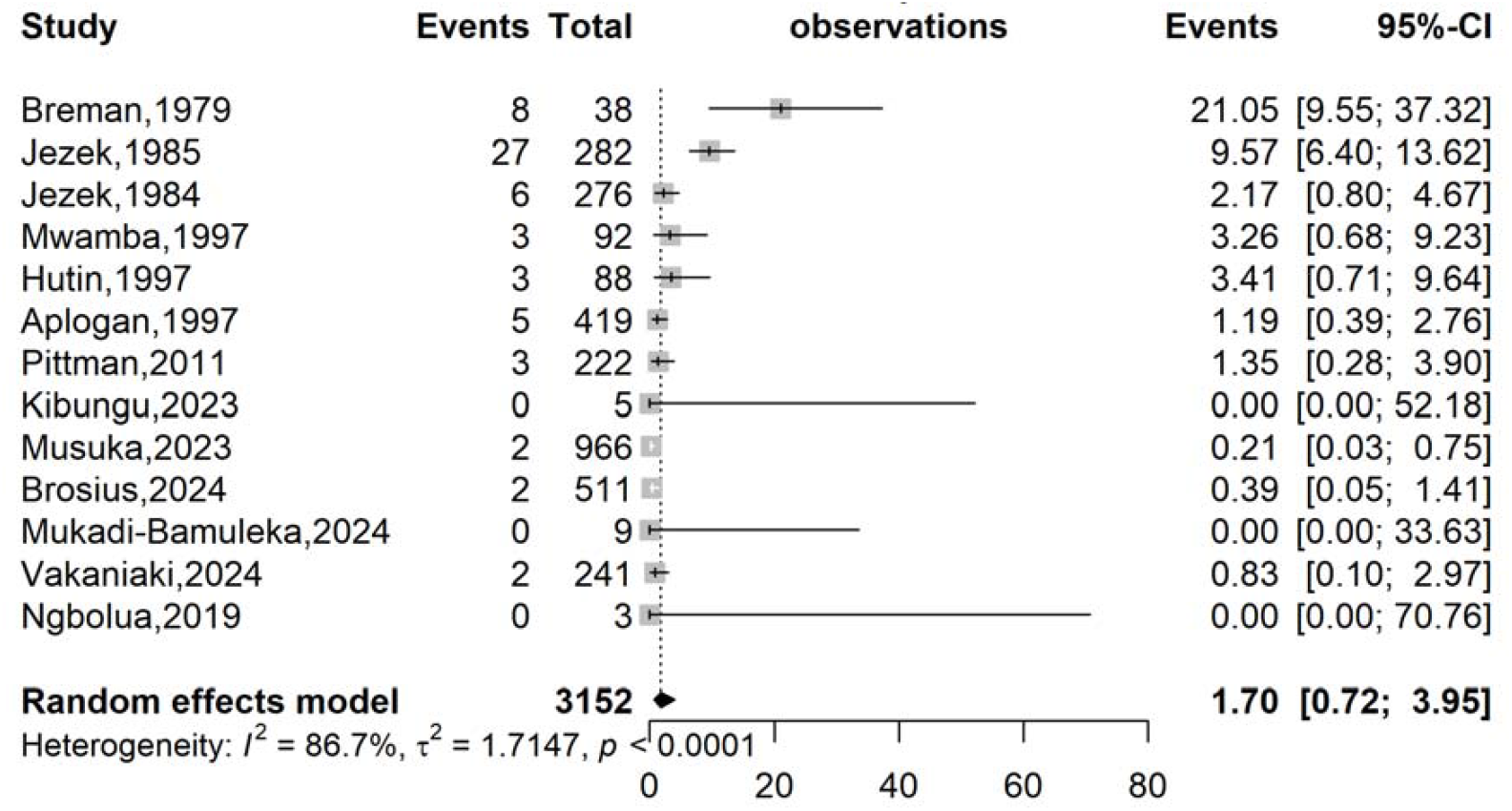
Pooled case fatality rate for suspected Mpox case in DRC, 1970-2024

### Mortality among confirmed Mpox cases

The pooled Case Fatality Rate (CFR) among confirmed Mpox cases was 2.46% (95% CI: 0.65-8.85; *I*^²^ = 90.3%, p < 0.001) (Fig. 4). Subgroup analysis revealed a significant decrease (*p* < 0.001) in CFR in the DRC from 7.29% (95% CI: 2.38-20.20; n = 5 studies) before 2022 to 0.28% (95% CI: 0.11-0.75; n = 4 studies) between 2011-2024. The highest burden of mortality was significantly observed (p < 0.001) in Western regions (13.25%; 95% CI: 13.25-34.34; n = 3 studies), while the lowest was in Eastern regions (0.45%; 95% CI: 0.11-1.80%; n = 2 studies). The CFR was higher within community settings (3.69%; 95% CI: 0.78-15.81; n = 7 studies) compared to hospital settings (0.77%; 95% CI: 0.32-1.84; n = 2 studies). Localities with the lowest burden of confirmed cases (< 200) presented with a higher CFR (19.12%; 95% CI: 11.44-30.20; n = 4 studies), and this was statistically significant (*p* < 0.001) (Table 3 and Additional Files 4, Supplementary Fig. 1-4)

**Table 3.**
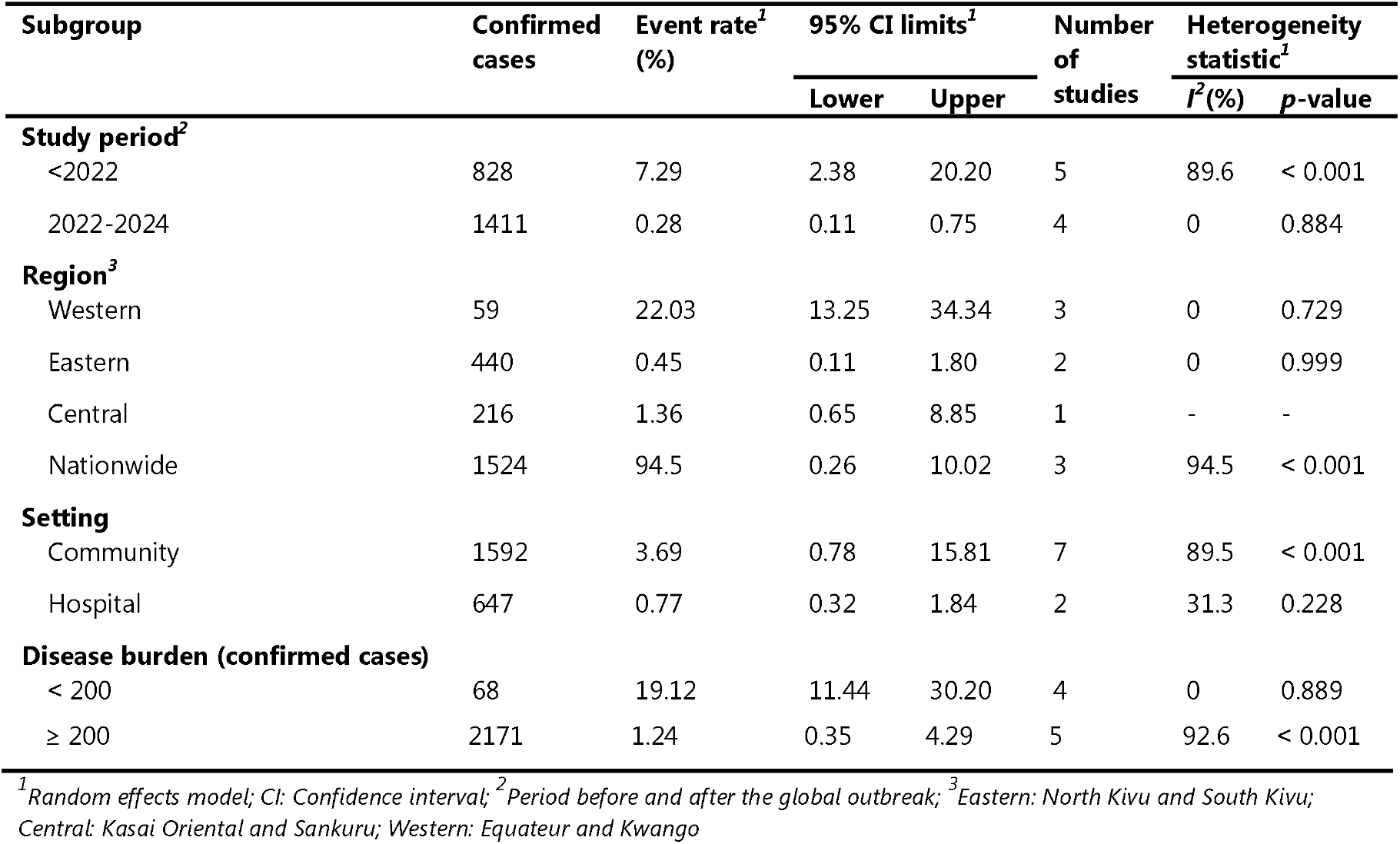
Subgroup meta-analysis of case fatality rate pooled estimates for confirmed Mpox cases in DRC, 1970-2024.

**Fig. 4.**
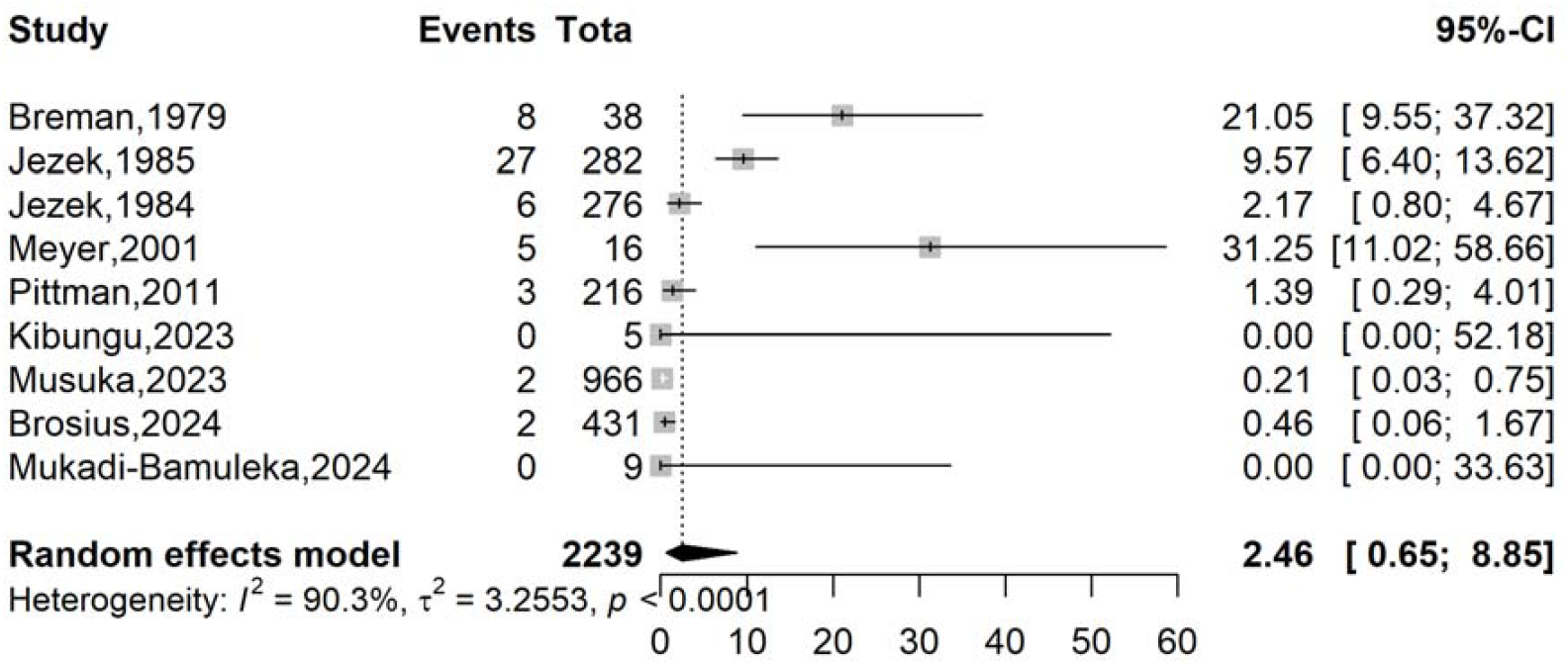
Pooled case fatality rate for confirmed Mpox case in DRC, 1970-2024

### Meta-regression analysis

Univariate analysis revealed that study year were significant sources of heterogeneity (*p* = 0.027) for the examined health indicators. In the multivariate analysis, study year, region and setting were significant sources of heterogeneity of suspected (*p* = 0.027) and confirmed case fatality rates (*p* = 0.016; *p* = 0.003; *p* = 0.041 respectively) (Table 4).

**Table 4.**
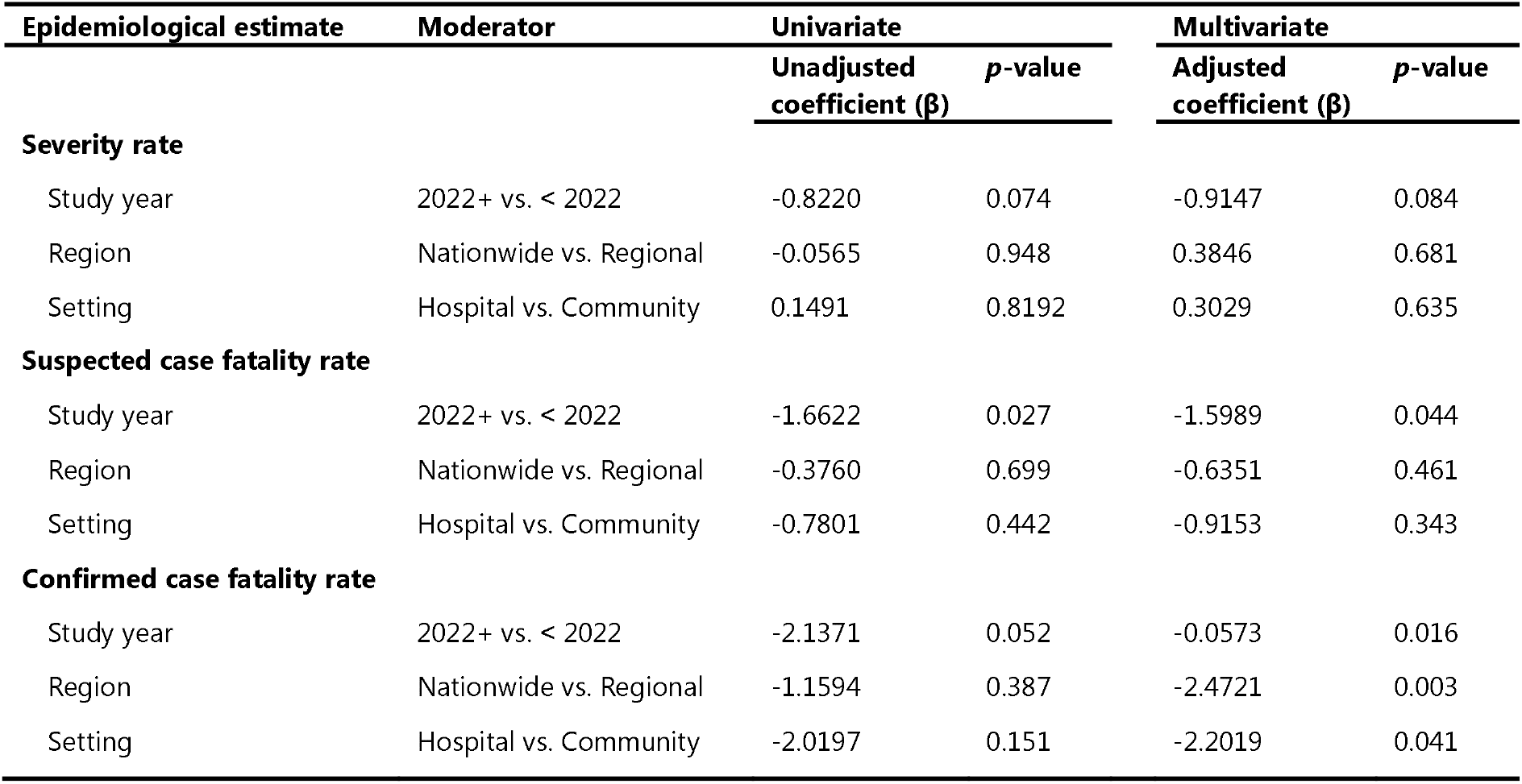
Univariate and multivariate meta-regression analysis of Mpox disease outcome metrics in DRC, 1970-2024.

### Publication bias and sensitivity test analysis

A large and almost symmetrical distribution of data points was observed in the funnel plot suggesting a low risk of publication bias for all the outcome interest. In addition, the Egger’s linear regression and Begg’s rank correlation tests confirmed the absence of statistically significant publication bias (Additional Files 2, Supplementary Fig. 6; Additional Files 3, Supplementary Fig. 6).

Sensitivity analysis, assessing the impact of individual studies and outliers on the overall results, demonstrated that no study exerted a significant impact on the overall pooled estimate (Additional Files 2, Supplementary Fig. 5; Additional Files 3, Supplementary Fig. 5, and Additional Files 4, Supplementary Fig. 5).

## Discussion

This systematic review and meta-analysis were conducted to determine the Mpox severity and CFRs in the world most endemic focus. It synthesized data from 19 studies to provide a comprehensive overview of Mpox severity and mortality in the DRC from 1970 to 2024.

### Severity rate

The observed pooled severity rate of 39.47% (95% CI: 29.11-50.86; *I*^*2*^ = 93.9% and *p*<0.001) highlights the substantial clinical burden of Mpox in the DRC. This finding is consistent with previous reports that have documented a range of severity in Mpox cases, with some individuals experiencing mild, self-limiting illness and others developing severe complications [1,8]. A study in the United Kingdom estimated the risk of hospitalization among 90,439 laboratory-confirmed Mpox cases to be between 3.53% (95% CI: 2.20-5.60) and 9.43% (95% CI: 5.18-16.57) [35]. This was lower than the proportion observed in our study. Factors such as host immunity, viral strain, and access to care probably influence the observed differences in disease severity [30]. Evidence indicates that infections with viral clade 1a result in more severe disease than infections with other clades [3].

A notable trend observed in the subgroup analysis was the progressive decline in Mpox severity rate over time, from 46.34% before 2022 to 25.83% between 2022-2024. This decline may reflect potential changes in the circulating viral strains or improvements in healthcare infrastructure, increased surveillance and reporting. Current evidence highlights the identification in 2023 of a newly discovered clade 1b through genomic analysis of strains from previously non-endemic provinces in the Eastern DRC. This clade might exhibit less virulence than the classical clade 1a strain [36]. However, it is important to interpret this trend with caution, as heterogeneity across studies and variations in case definitions and methodology could also influence these findings.

Regional variations in Mpox severity were also evident, with the highest severity rate significantly observed in the Western regions (43.82%; *p* < 0.001) and suggest the influence of region-specific factors on disease presentation and outcomes. The Western DRC is endemic for the Congo Basin clade of Mpox virus, which has been associated with higher virulence and more severe clinical outcomes compared to the West African clade [37,38]. The existence of comorbidities like HIV could exacerbate Mpox severity, as underscored by a study in Nigeria and other African countries where the high prevalence and non-suppression of HIV/AIDS increases the risk of severe outcomes for Clade 1 Mpox virus infection [39,40]. Further research is needed to explore these regional disparities and identify potential risk factors for severe Mpox in specific areas.

To effectively address the ongoing Mpox threat, a multi-faceted approach is essential. Community-based interventions, including the deployment of mobile health units, implementation of community education programs, and provision of early diagnostic tools, are crucial to reducing delays in care-seeking and improving outcomes, particularly given the observed high severity of Mpox in community settings.

### Case fatality rate

The pooled CFR among suspected Mpox cases was 1.70% (95% CI: 0.72-3.95; *I*^2^ = 86.7% and *p*<0.001), with a significant decreasing trend observed over time (*p* < 0.001). This finding aligns with previous studies that have reported CFRs for Mpox generally ranging from 0% to 12% [3]. The pooled CFR for confirmed Mpox cases in this analysis was 2.46% (95% CI: 0.65–8.85), exhibiting significant heterogeneity (*I*^²^ = 90.3%, *p* < 0.001). This result corroborates findings reported in an African study, where 45,652 Mpox cases, clinically diagnosed and laboratory-confirmed across 12 African countries, resulted in 1492 deaths (CFR, 3.3%) [41]. The high heterogeneity suggests substantial variability in CFR across the included studies, which can likely be attributed to several factors, including temporal shifts, regional disparities, and differences in healthcare settings.

A notable trend observed was the decline in CFR over time. The CFR was considerably higher (7.29%) in studies conducted before 2022, likely reflecting the limited healthcare access and absence of Mpox-specific interventions during that period. The cessation of smallpox vaccination campaigns around 1980 may have also contributed to increased susceptibility to Mpox, particularly in older cohorts [15]. In contrast, the CFR decreased significantly to 0.28% in studies conducted between 2022 and 2024. This decline aligns with improvements in surveillance, earlier case detection, and enhanced supportive care [32]. However, it is important to acknowledge that potential underreporting of milder Mpox cases in recent years could introduce a downward bias in the CFR estimates.

Regional disparities also played a significant role in the observed heterogeneity (p = 0.003). The Western regions of the DRC exhibited a substantially higher CFR (22.03%) compared to the Eastern regions (0.45%). This difference may be attributed to the predominance of the more virulent Congo Basin clade in the Western/Central DRC and the presence of rural healthcare gaps that contribute to delays in seeking and receiving appropriate treatment. In contrast, the Eastern DRC generally has better urban healthcare infrastructure, which may contribute to improved outcomes. Additionally, the possibility of cross-protection from Ebola vaccination campaigns in the Eastern DRC as a contributing factor to the lower CFR in that region should be also explored [42,43].

Furthermore, the healthcare setting influenced the CFR (p = 0.041). Community settings were associated with a higher CFR (3.69%) compared to hospital settings (0.77%). This finding likely reflects factors such as late presentation, misdiagnosis, and the use of traditional medicine in community settings, which can exacerbate disease outcomes. Study reports establish that traditional African Medicine remains an important health care resource among population in DRC [44]. On the other hand, hospital settings offer access to supportive care, such as fluid management and antibiotics for secondary bacterial infections, which can reduce mortality.

Localities with a high burden of confirmed Mpox cases (≥ 200 cases) demonstrated a lower CFR (1.24%). This finding may be explained by enhanced awareness leading to earlier case identification and reporting, as well as more efficient resource allocation and targeted interventions, such as vaccination and healthcare worker training, in high-burden areas [32]. The findings of this review have several implications for research and public health policy. There is a clear need for targeted interventions in the Western and Central DRC, including healthcare strengthening initiatives such as mobile clinics to reach remote populations and the potential use of strain-specific vaccines given the virulence of the Congo Basin clade.

## Limitations and strength

This review has several limitations. First, some death cases might have been unreported due to study design, potentially underestimating the true CFR. Confounding by strain differences remains a concern, as the lack of comprehensive genomic data in many studies obscures the clade-specific mortality rates. Surveillance bias, particularly the potential for improved detection of milder cases in more recent studies, may also influence the CFR estimates. Additionally, the small sample sizes in some subgroup analyses result in wide confidence intervals, therefore limiting the precision of the findings. Study in other language than English and French were not searched. Despite these limitations, this systematic review and meta-analysis provides valuable insights into Mpox severity and mortality in the DRC. The findings highlight the continued public health importance of Mpox in the region and underscore the need for ongoing surveillance, research, and interventions to reduce the burden of this disease

## Conclusions

This systematic review and meta-analysis provide a comprehensive assessment of Mpox severity and mortality in the DRC from 1970 to 2024. The pooled analyses revealed an overall Mpox severity rate of 39.47% and a Case Fatality Rate (CFR) of 2.46% among suspected Mpox cases. Subgroup analyses and meta-regression identified significant temporal, regional and study setting variations, highlighting the complex and evolving epidemiology of Mpox in the DRC. Despite the limitations inherent in the available data, this study underscores the continued public health burden of Mpox in the DRC and emphasizes the need for sustained surveillance, research, and interventions aimed at ensuring continuous vaccines supply, community engagement, and healthcare system strengthening.

## Supporting information

Addition Files 2

Addition Files 3

Addition Files 4

## Data Availability

All data produced in the present work are contained in the manuscript

## Abbreviations

CFR: Case Fatality Rate
CI: Confidence Interval
DRC: Democratic Republic of Congo
HCW: Healthcare worker
HIV/AIDS: Human Immunodeficiency Virus/Acquired Immunodeficiency Syndrome
MeSH: Medical Subject Headings
PCR: Polymerase Chain Reaction
PRISMA: Preferred Reporting Items for Systematic Reviews and Meta-Analysis

## Declarations

### Author contributions

F.Z.L.C. conceived the original idea of the study. F.Z.L.C. and A.Z. conducted the literature search. F.Z.L.C., A.Z. and S.N. selected the studies, extracted the relevant information, and synthesized the data. F.Z.L.C. performed the analyses and wrote the first draft of the manuscript. All authors critically reviewed and revised successive drafts of the manuscript. All authors read and approved the final manuscript.

### Ethical approval statement

Not applicable

### Consent for publication

Not applicable.

### Availability of data and materials

The sources of data supporting this systematic review are available in the reference. All data generated or analyzed during this study are included in this published article and supplemental material.

