## Additional Files 1 for "Mpox severity and mortality in the DRC: a systematic review and meta-analysis (1970-2024)"

Supplementary Table 1: Characteristic of studies assessing Mpox severity and mortality in DRC, 1970-2024

| **Author** | **Study**  **Year*^1^*** | **Region** | **Setting** | **Study population** | **Study type** | **Sampling** | **Risk of bias** | **Severity rate (%)** | **CFR rate**  **(%)** | **Summary of findings** |
| --- | --- | --- | --- | --- | --- | --- | --- | --- | --- | --- |
| Breman *et al.* [1] | 1979 | Equateur | Community | General population | Cross-sectional | Non-probabilistic | Low | 57.89 | S: 21.05  C: 21.05 | Of the 47 reported cases in Central and West Africa, 38, representing the vast majority, occurred in Zaire. These cases, largely in children under 10, presented with a 17% case-fatality rate. |
| Jezek *et al.* [2] | 1984 | Nationwide | Community | General population | Cross-sectional | Non-probabilistic | Low | 39.01 | S: 2.17  C: 2.17 | A study of 2,510 contacts of monkeypox patients revealed a significant number of secondary cases, primarily in children, with waning immunity observed in vaccinated individuals, though vaccination still offered considerable protection. |
| Jezek *et al.* [3] | 1985 | Nationwide | Community | General population | Cross-sectional | Non-probabilistic | Low | 68.75 | S: 9.57  C: 9.57 | Unvaccinated individuals, particularly young children, experienced an 11-15% case-fatality rate, while vaccinated individuals showed no deaths and altered disease presentation. |
| Mwamba *et al.* [4] | 1997 | Kasai Oriental | Community | General population | Cross-sectional | Non-probabilistic | Low | NR | S: 3.26  C: NR | The 1996-97 investigation suggested most monkeypox cases resulted from person-to-person transmission, a shift from earlier reports of primarily animal-to-human spread. |
| Hutin *et al.* [5] | 1997 | Sankuru | Community | General population | Cross-sectional | Non-probabilistic | Low | NR | S: 3.41  C: NR | This outbreak presented with notable attack and case-fatality rates. The cessation of smallpox vaccination in 1983, following global eradication, contributed to an increased population susceptibility to monkeypox. |
| Aplogan *et al.* [6] | 1997 | Kasai Oriental | Community | General population | Cross-sectional | Non-probabilistic | Low | NR | S: 1.19  C: NR | There was a high prevalence of Mpox among children and a mix of primary and secondary transmission patterns across numerous villages. The outbreak was localized. Travel and close contact within households and neighborhoods facilitated the secondary case transmission. |
| Meyer *et al.* [7] | 2001 | Equateur | Community | General population | Cross-sectional | Non-probabilistic | Low | 68.75 | S: NR  C: 31.25 | Two outbreaks were confirmed as monkeypox (4 deaths), two as co-infection of monkeypox and chickenpox (1 death), two as chickenpox (no deaths), and one outbreak yielded no viral evidence (no deaths). |
| Pittman *et al.* [8] | 2022 | Sankuru | Hospital | General population | Cross-sectional | Non-probabilistic | Moderate | 63.89 | S: 1.35  C: 1.39 | Clinical course of Mpox in 216 PCR-confirmed cases revealed a 1.4% mortality rate and significant fetal loss in pregnant patients. Key findings included a high prevalence of rash and lymphadenopathy, with younger children exhibiting higher lesion counts and severe disease being associated with hypoalbuminemia and elevated viral load. |
| Hughes *et al.* [9] | 2014 | Tshuapa | Community | General population | Cross-sectional | Non-probabilistic | Low | 36.52 | NR | A significant proportion of mpox cases were co-infected with varicella zoster virus (VZV), exhibiting atypical clinical presentations and highlighting the complex interplay between these two viruses. |
| Osadebe *et al.* [10] | 2014 | Tshuapa | Community | General population | Cross-sectional | Non-probabilistic | Low | 28.53 | NR | Laboratory-confirmed Mpox and VZV cases presented with many of the same signs and symptoms, and the analysis here emphasized the utility of including 12 specific signs/symptoms when investigating Mpox cases |
| Whitehouse *et al.* [11] | 2014 | Tshuapa | Community | General population | Cross-sectional | Non-probabilistic | Low | 50.43 | NR | Increased incidence compared to previous decades, likely due to waning smallpox immunity. While males generally had higher infection rates, females reported frequent contact with symptomatic individuals. Animal exposures were most common in males. |
| Mande *et al.* [12] | 2019 | Bas-Uélé | Community | General population | Cross-sectional | Non-probabilistic | Low | 38.10 | S: NR  C: NR | Among 77 suspected cases, PCR revealed 27.3% monkeypox, 58.4% chickenpox, and 14.3% negative. Monkeypox cases showed distinct skin lesions. |
| Ngbolua et al. [13] | 2019 | North Ubangui | Hospital | General population | Case report | Non-probabilistic | High | NR | S: 0.00  C: NR | Three cases of monkeypox in young males with similar clinical presentations, including fever, rash, itching, and abdominal pain. No case of death. |
| Kibungu *et al.* [14] | 2023 | Kwango | Community | General population | Case report | Non-probabilistic | Low | 0.00 | S: 0.00  C: 0.00 | A cluster of clades I monkeypox cases in the DRC shows sexual transmission, indicating this route is not limited to clade IIb. |
| Musuka *et al.* [15] | 2023 | Nationwide | Community | General population | Cross-sectional | Non-probabilistic | Low | NR | S: 0.21  C: 0.21 | The Mpox prevalence in Central Africa is linked to bushmeat, lack of smallpox vaccination, HIV, and close contact. Clinical features included rash, fever, and lymphadenopathy. |
| Brosius *et al.* [16] | 2024 | South Kivu | Hospital | General population | Cross-sectional | Non-probabilistic | Moderate | 24.83 | S: 0.39  C: 0.46 | Most suspected cases were PCR-positive for Mpox. Most cases reported contact with known Mpox, primarily spouses/partners in adults and family in children. Genital lesions were common in adults. Hospitalized mortality was low. |
| Mukadi-Bamuleka e*t al.* [17] | 2024 | North Kivu | Community | General population | Case report | Non-probabilistic | Low | 66.67 | S: 0.00  C: 0.00 | Clade Ib monkeypox was introduced into North Kivu, including displacement camps, with suspected non-intimate contact transmission, and affecting children. |
| Kombozi *et al.* [18] | 2024 | Tshopo | Community | General population | Cross-sectional | Probabilistic | Low | 44.35 | NR | Occupational activities, such as hunting and agriculture, alongside economic vulnerability and direct exposure to infected animals or individuals, play a significant role in Mpox transmission. These results underscore the complex interplay between human behavior, environmental interaction, and disease spread. |
| Vakaniaki *et al*. [19] | 2024 | South Kivu | Community | General population | Cross-sectional | Non-probabilistic | Low | 9.26 | S: 0.83  C: NR | The Mpox outbreak in eastern DRC was caused by a distinct Clade I Mpox lineage, differing from historical zoonotic patterns. The outbreak, predominantly affected young adults including a significant proportion of female sex workers, suggests a shift towards human-to-human transmission, potentially involving sexual contact. |
| *^1^ Date of study completion; CFR: Case Fatality Rate; VZV: Varicella-Zoster Virus; S: Among suspected Mpox case; C: Among confirmed Mpox cases; NR: Not Reported; DRC: Democratic Republic of Congo; PCR: Polymerase Chain Reaction; HIV: Human Immunodeficiency Virus* | | | | | | | | | | |

Supplementary Table 2 Searching strategies for online databases

| Database | Search Term | Results |
| --- | --- | --- |
| PubMed | (  ("monkeypox"[MeSH Terms] OR "monkeypox"[Title/Abstract] OR "mpox"[Title/Abstract] OR "MPXV"[Title/Abstract] OR "monkeypox virus"[MeSH Terms])  AND  ("severity"[Title/Abstract] OR "severity"[MeSH Terms] OR "surveillance"[MeSH Terms] OR "surveillance"[Title/Abstract] OR "mortality"[Title/Abstract] OR "mortality"[MeSH Terms] OR "death"[Title/Abstract] OR "death"[MeSH Terms] OR "fatality"[Title/Abstract] OR "case fatality"[Title/Abstract])  AND  ("Democratic Republic of the Congo"[MeSH Terms] OR "Democratic Republic of Congo"[Title/Abstract] OR "DRC"[Title/Abstract] OR "Congo"[Title/Abstract] OR "Zaire"[Title/Abstract])  ) | 153 |
| Web of Science | ("monkeypox" OR "mpox" OR "MPXV" OR "monkeypox virus")  AND  ("severity" OR "surveillance" OR "mortality" OR "death" OR "fatality" OR "case fatality")  AND  ("Democratic Republic of Congo" OR "DRC" OR "Congo" OR "Zaire") | 283 |
| ScienceDirect | ("monkeypox" OR "mpox) AND ("severity" OR “surveillance” OR "mortality" OR "death") AND (Democratic Republic of Congo" OR DRC ) | 820 |
| Scopus | (TITLE-ABS-KEY((monkeypox OR "monkeypox virus" OR "human monkeypox" OR Mpox OR mpox OR MPX)  AND  (epidemiology OR surveillance OR characteristics OR "clinical characteristics" OR severe OR mortality OR death)  AND  (DRC OR "Democratic Republic of Congo" OR Zaire)) | 140 |
| Embase | ( 'monkeypox'/exp OR 'monkeypox':ti,ab )  AND  ( 'severity':ti,ab OR 'mortality':ti,ab )  AND  ( 'Democratic Republic of the Congo'/exp OR 'DRC':ti,ab ) | 25 |
| CINAHL | ( TI ("monkeypox" OR "mpox") OR SU "Monkeypox" )  AND  ( TI ("severity" OR "mortality") OR SU "Severity of Illness" OR SU "Mortality" )  AND  ( TI ("Democratic Republic of Congo" OR "DRC") OR SU "Democratic Republic of the Congo" ) | 4 |
| AJOL | “Monkeypox” or “Mpox” and “Democratic Republic of Congo" OR "DRC" | 4013 |
