## Supplementary material for "Mpox severity and mortality in the DRC: a systematic review and meta-analysis (1970-2024)": Addition Files 2

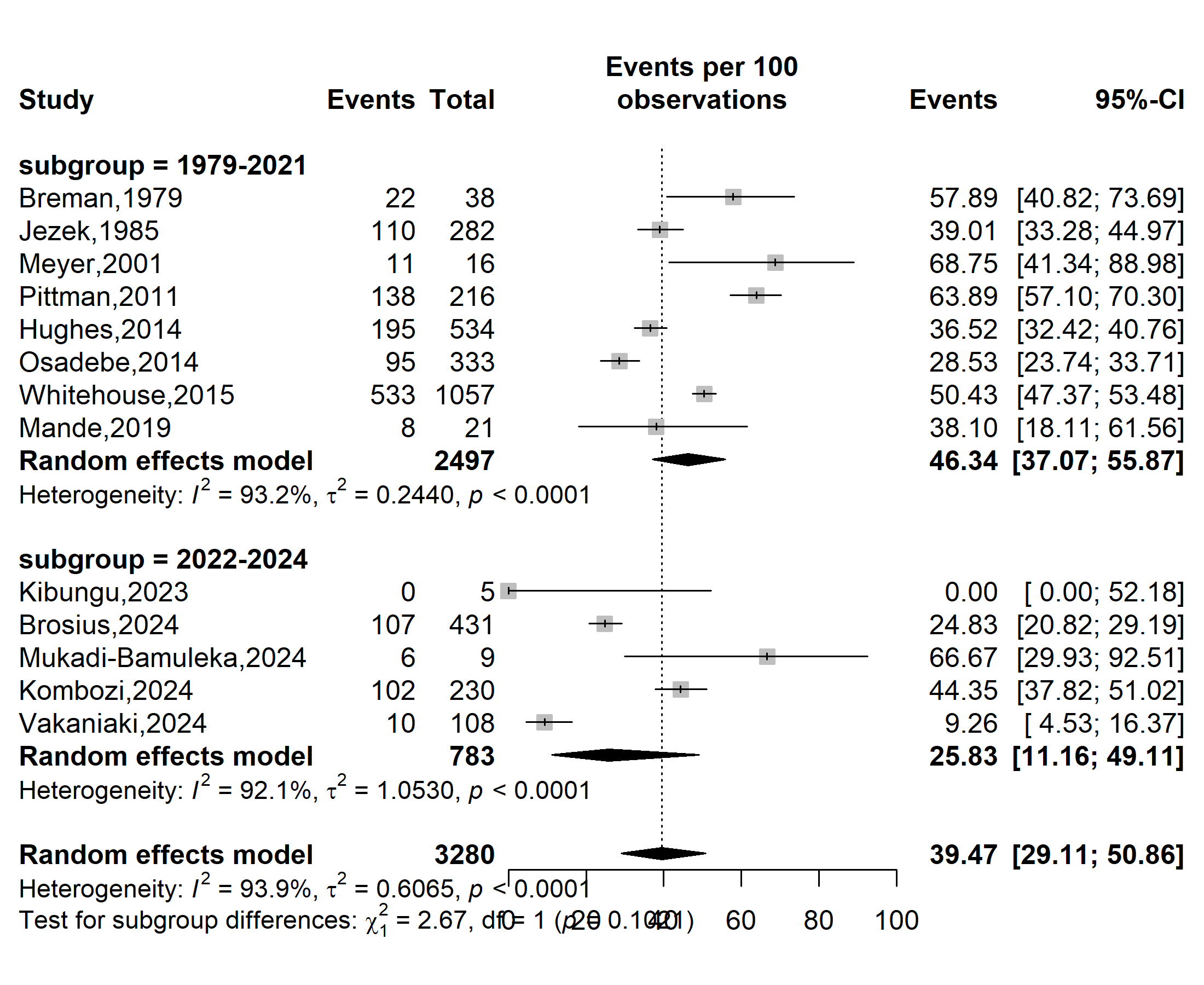

**Severity rate (%)**

**Event rate (%)**

Supplementary Fig. 1 Subgroup estimates of the Mpox severity rate in DRC, 1970-2024

*(by before and after the global Mpox outbreak)*

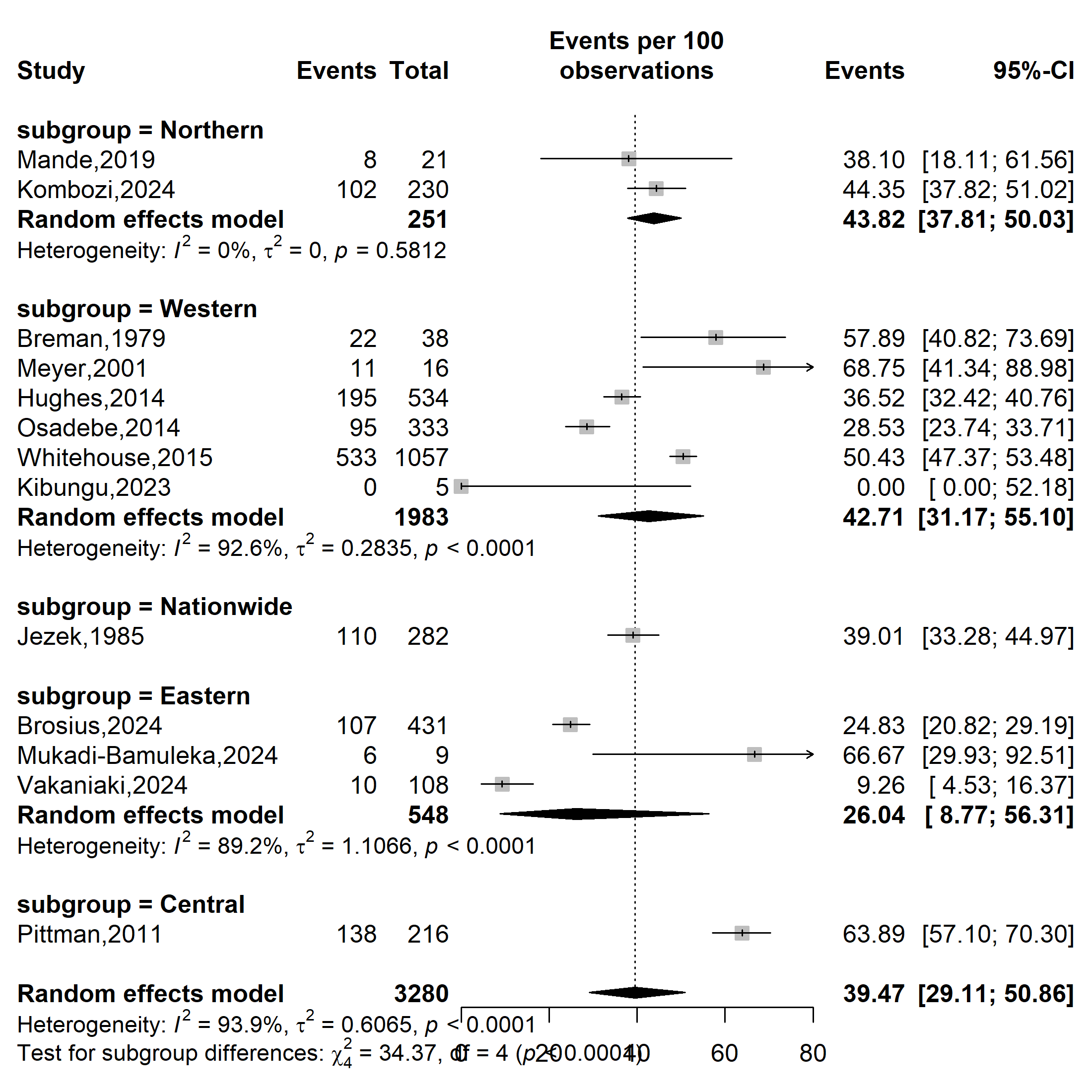

**Severity rate (%)**

**Event rate (%)**

Supplementary Fig. 2 Subgroup estimates of the Mpox severity rate in DRC, 1970-2024

*(based on geographical localization: Eastern: North and South Kivu; Northern:* *Bas-uélé and Tshopo; Western: Equateur, Tshuapa, and Kwango; Central: Sankuru)*

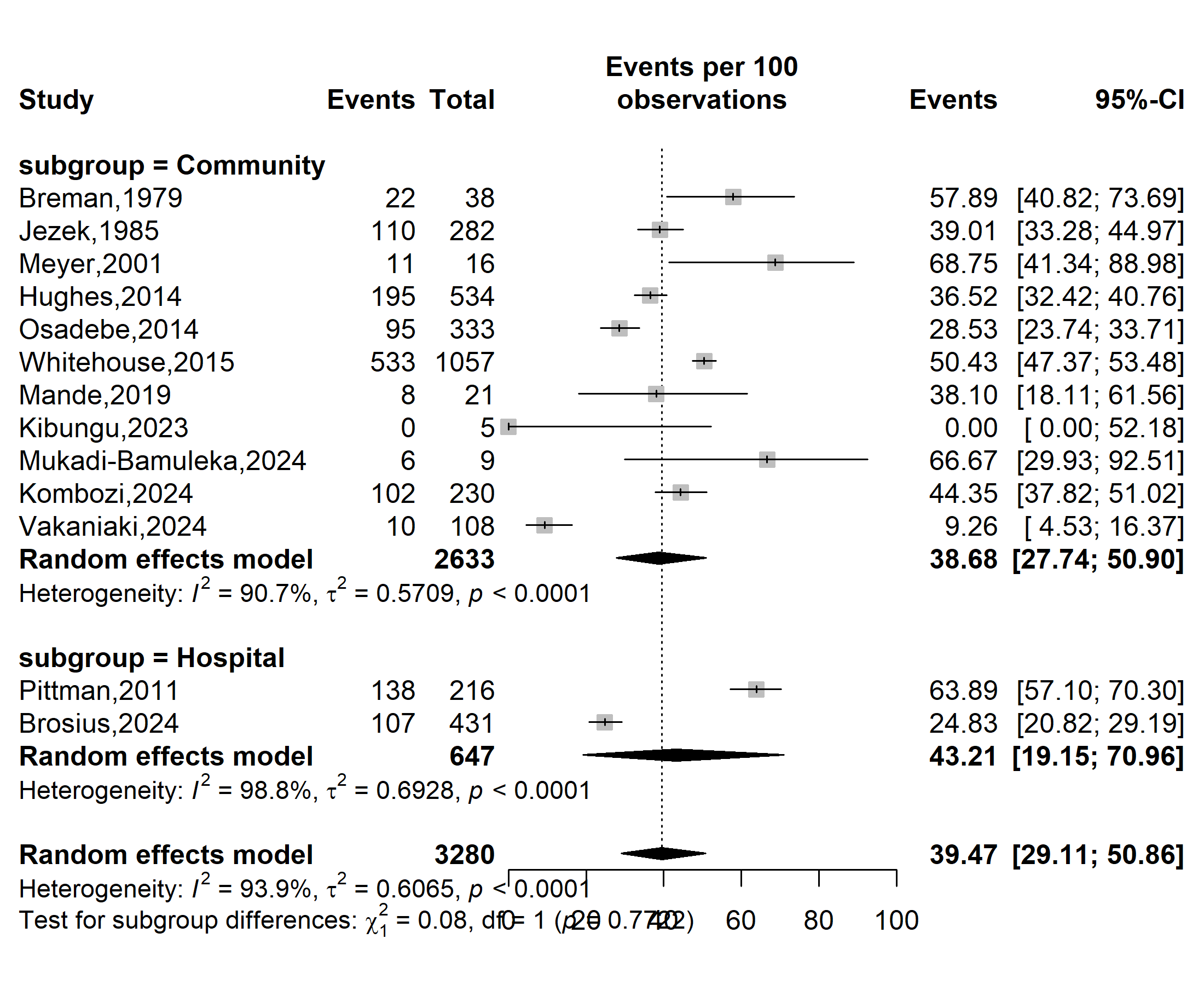

**Event rate (%)**

**Severity rate (%)**

Supplementary Fig. 3 Subgroup estimates of the Mpox severity rate in DRC, 1970-2024

*(based on study setting)*

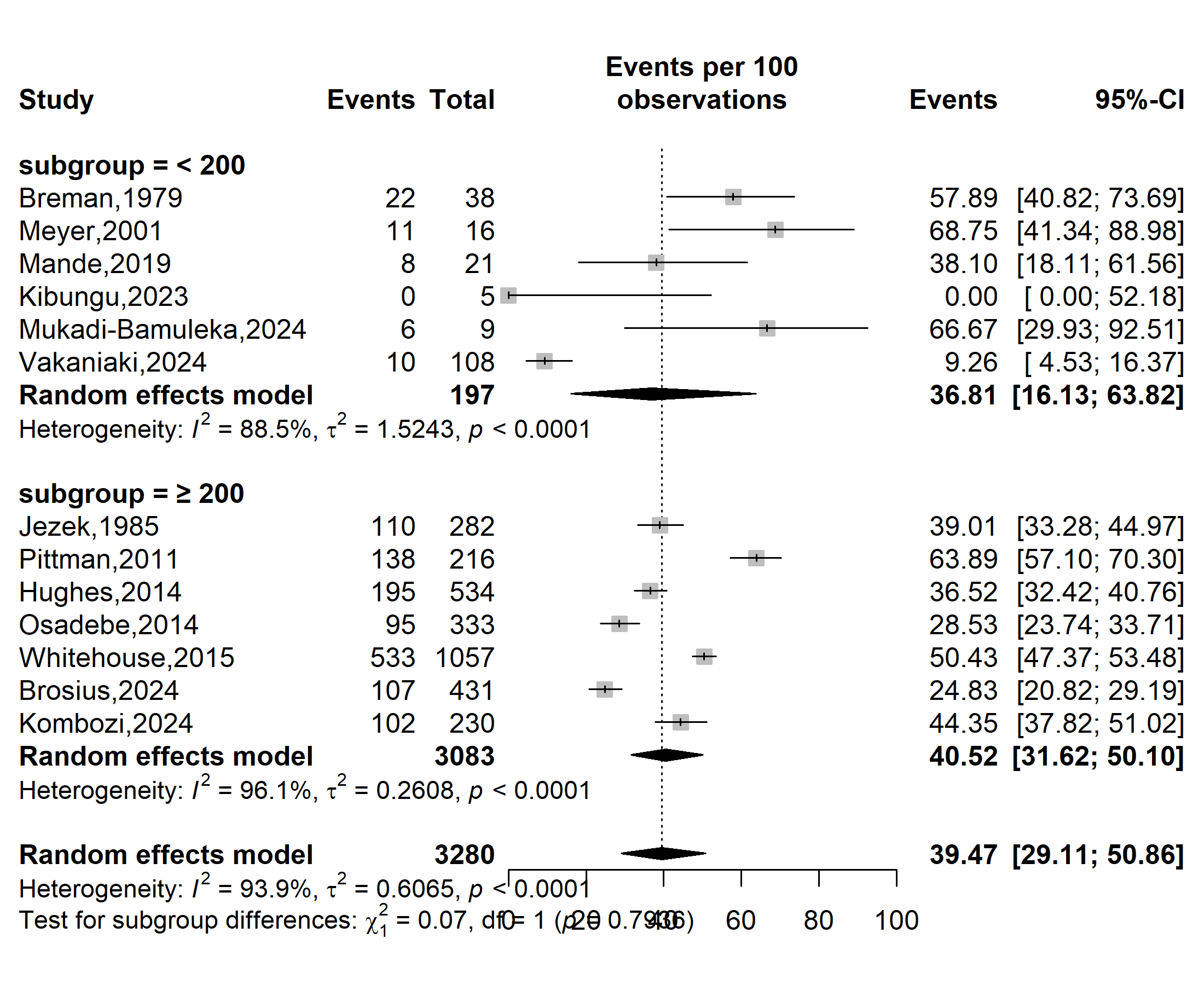

**Event rate (%)**

**Severity rate (%)**

Supplementary Fig. 4 Subgroup estimates of the Mpox severity rate in DRC, 1970-2024

*(Based on the disease burden: median number of confirmed cases)*

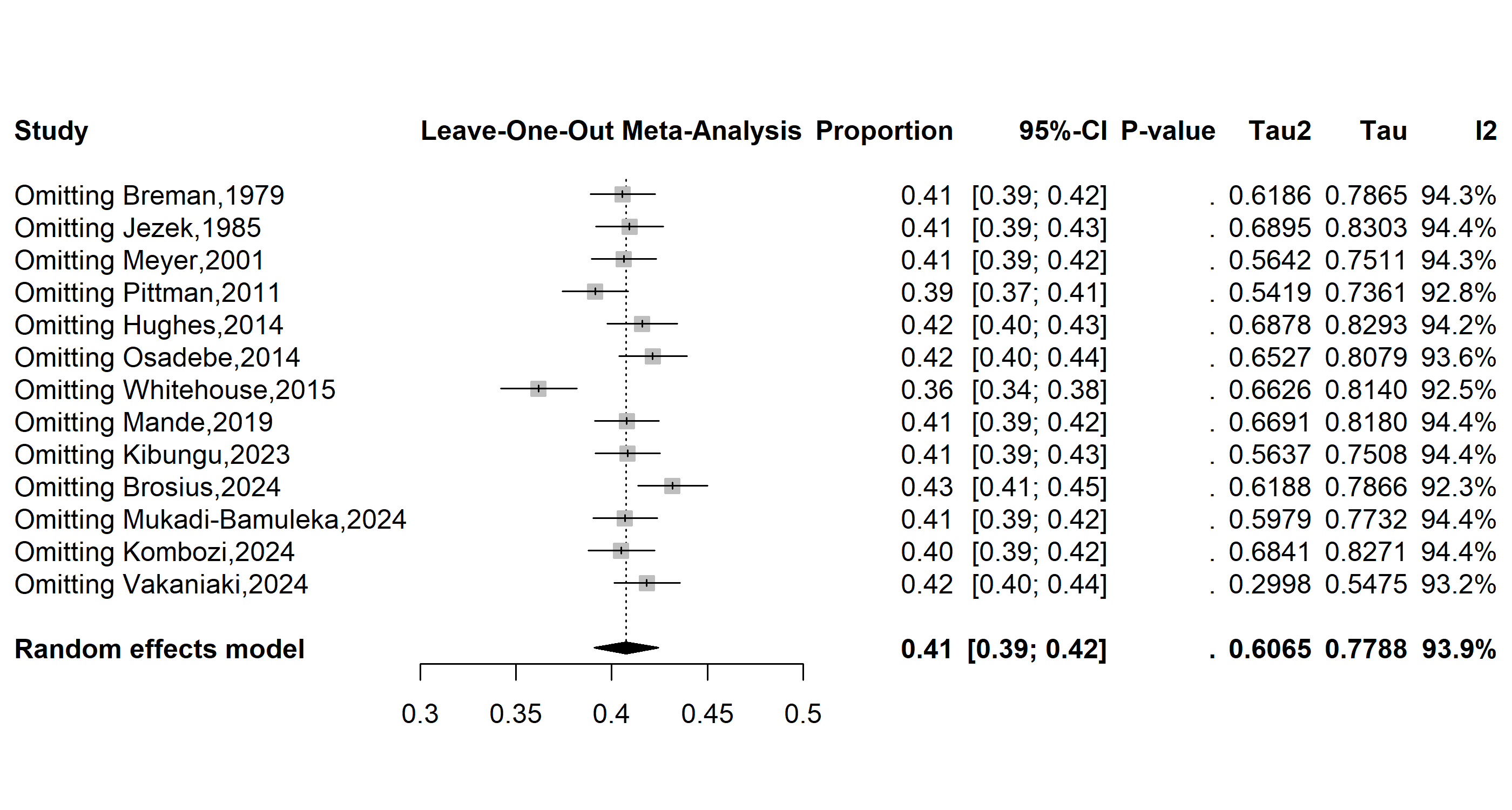

Supplementary Fig. 5 Sensitivity analysis of the pooled Mpox severity rate estimates in DRC, 1970-2024

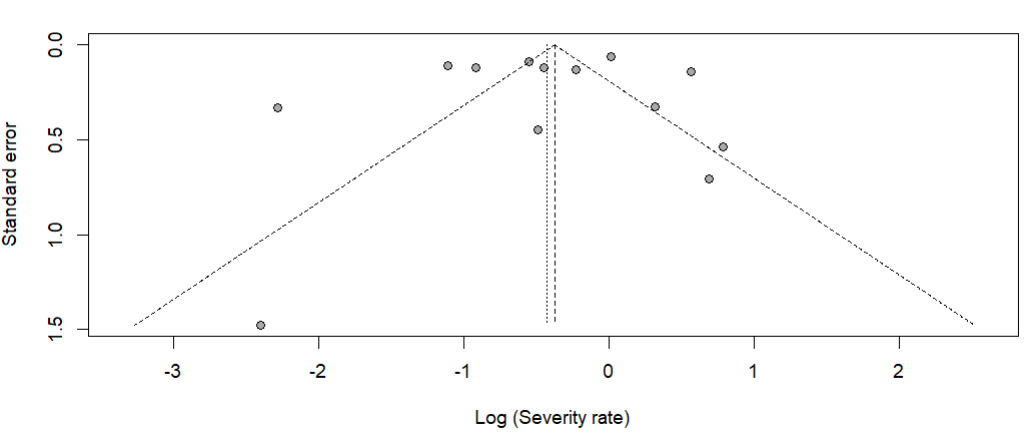

Egger’s test, *p*-value = 0.638

Begg’s test, *p*-value = 0.626

Supplementary Fig. 6 Funnel plot with pseudo 95% confidence limits and tests assessing the publication bias studies included
