## Supplementary material for "Mpox severity and mortality in the DRC: a systematic review and meta-analysis (1970-2024)": Addition Files 3

**Event rate (%)**

**Case fatality rate (%)**

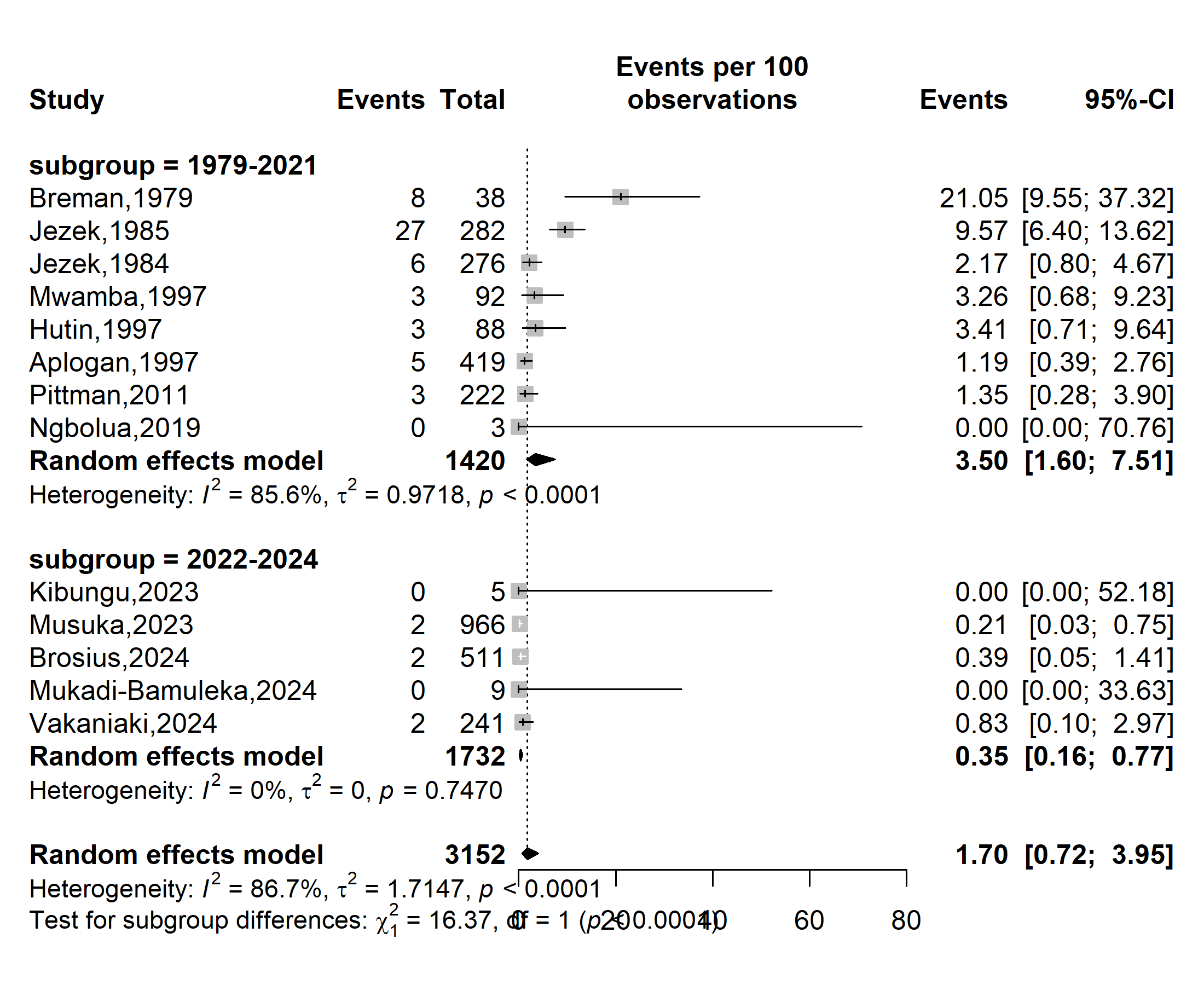

Supplementary Fig. 1 Subgroup estimates of the case fatality rate for suspected Mpox cases in DRC, 1970-2024 *(based on the period before and after the global outbreak)*

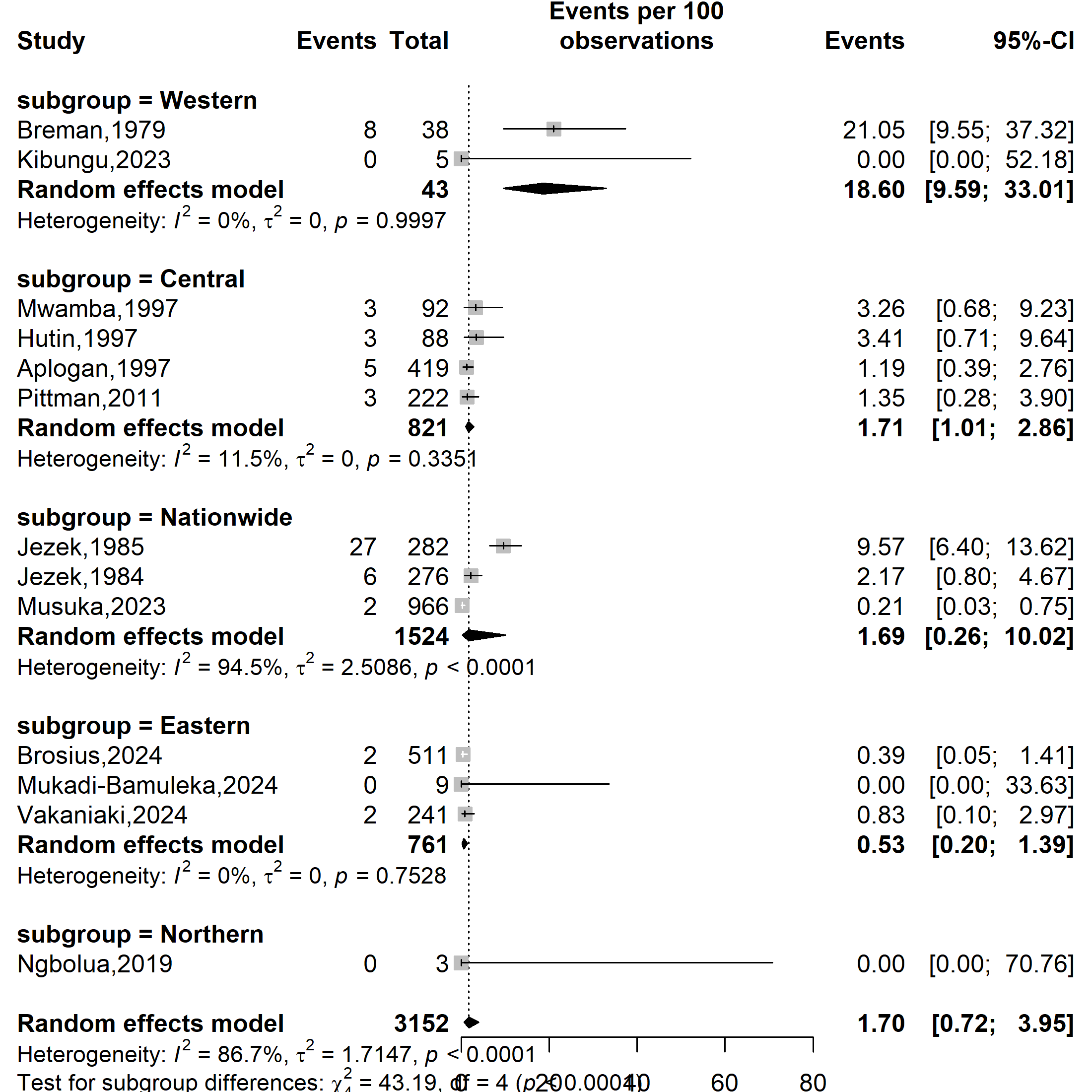

**Case fatality rate (%)**

**Event rate (%)**

Supplementary Fig. 2 Subgroup estimates of the case fatality rate for suspected Mpox cases in DRC, 1970-2024 *(based on geographical localization: Eastern: North Kivu and South Kivu; Central: Kasai Oriental and Sankuru; Northern: North Ubangui; Western: Equateur and Kwango)*

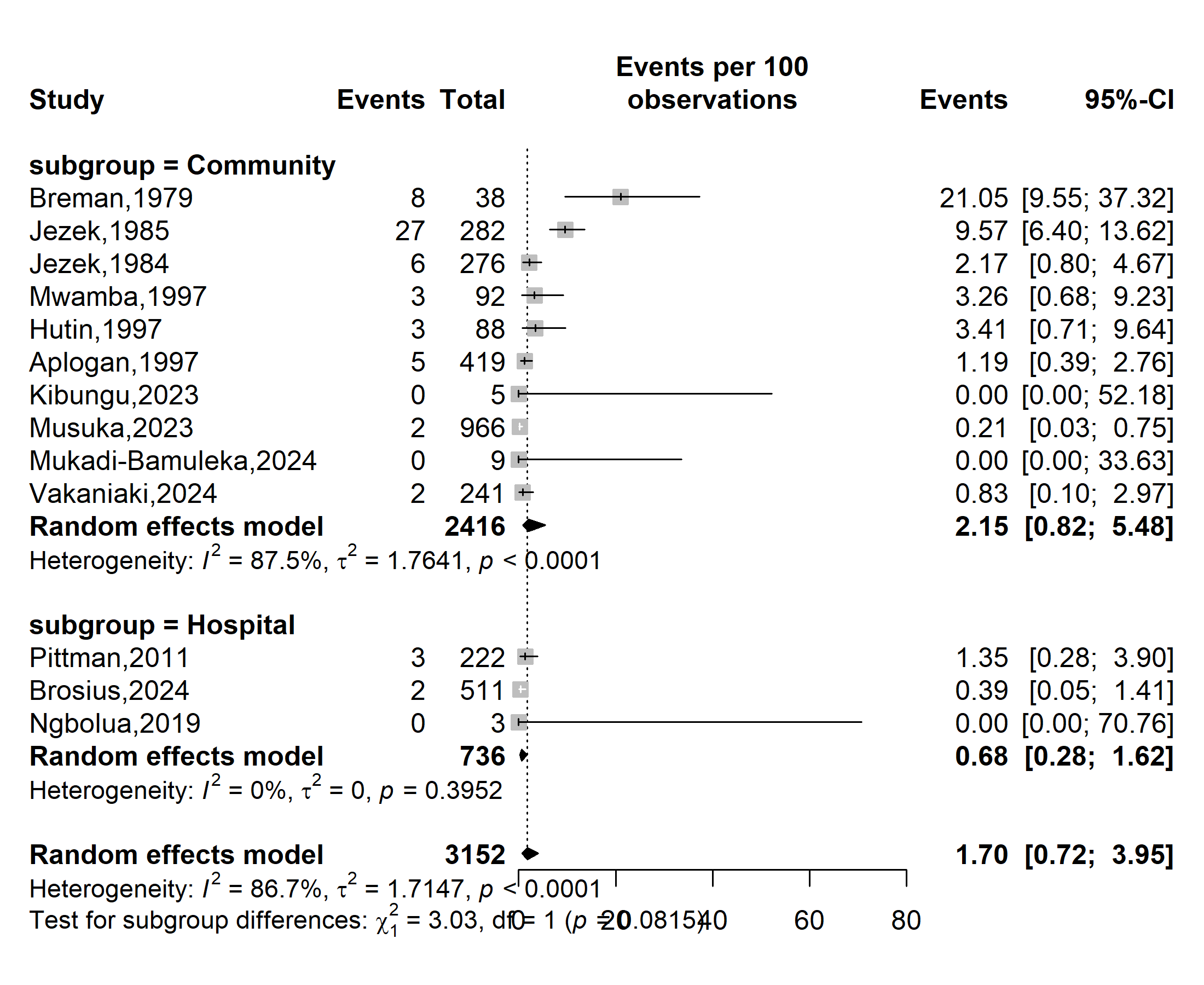

**Case fatality rate (%)**

**Event rate (%)**

Supplementary Fig. 3 Subgroup estimates of the case fatality rate for suspected Mpox cases in DRC, 1970-2024 *(based on study setting)*

**Case fatality rate (%)**

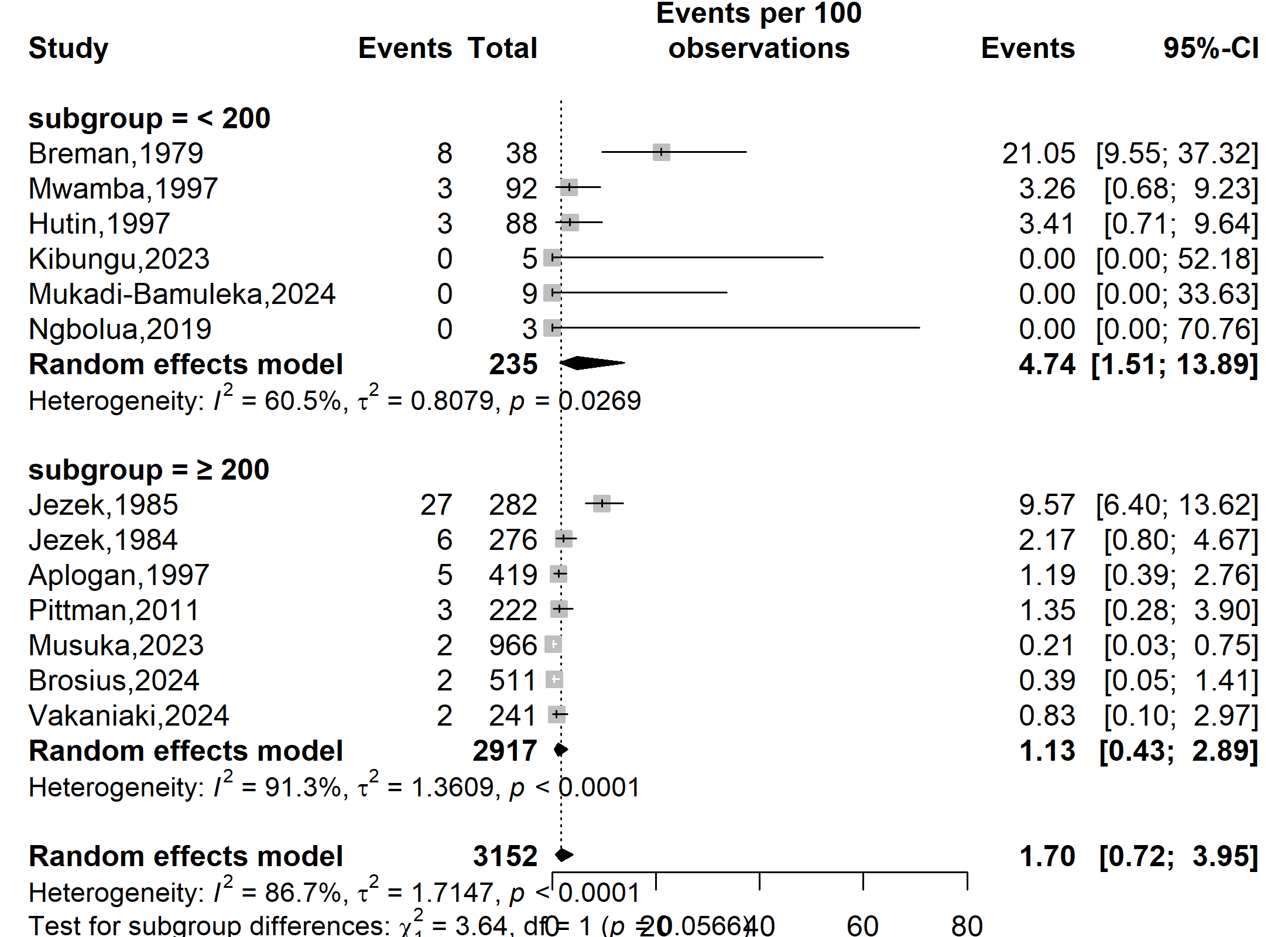

**Event rate (%)**

**Case fatality rate (%)**

Supplementary Fig. 4: Subgroup estimates of the case fatality rate for suspected Mpox cases in DRC, 1970-2024 *(based on the burden of disease: number of suspected cases)*

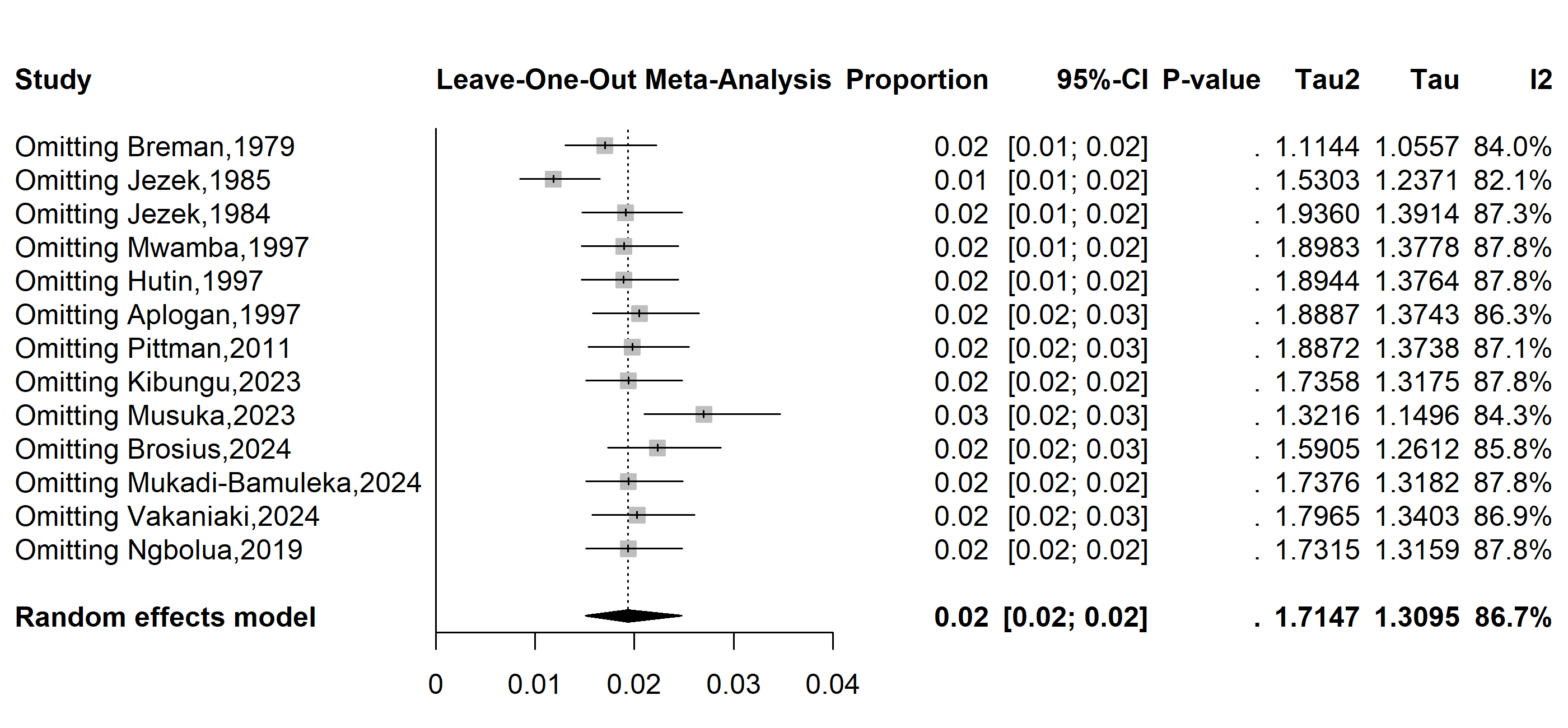

Supplementary Fig. 5 Sensitivity analysis of the pooled case fatality rate for suspected Mpox cases in DRC, 1970-2024

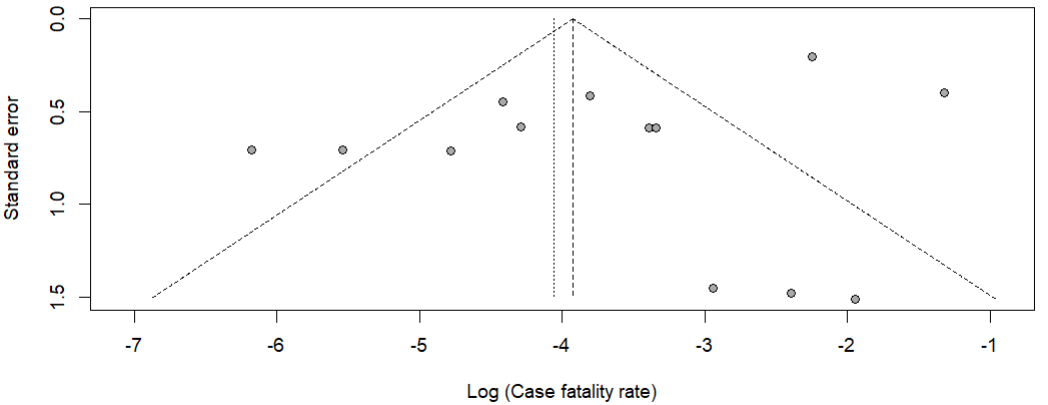

Egger’s test, *p*-value = 0.078

Begg’s test, *p*-value = 1.000

Supplementary Fig. 6 Funnel plot with pseudo 95% confidence limits and tests assessing the publication bias studies included
