## Supplementary material for "Mpox severity and mortality in the DRC: a systematic review and meta-analysis (1970-2024)": Addition Files 4

**Case fatality rate (%)**

**Event rate (%)**

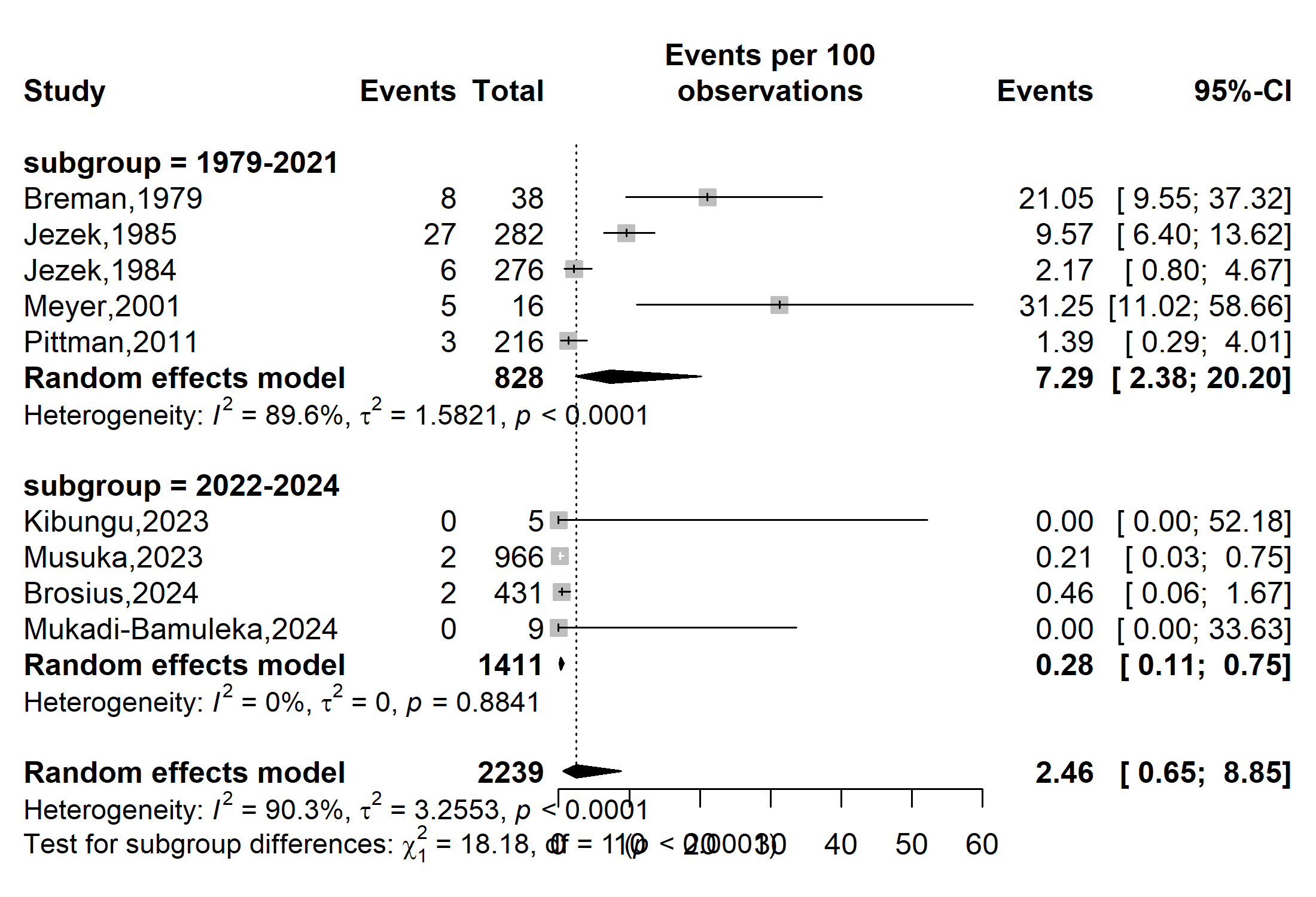

Supplementary Fig. 1 Subgroup estimates of the confirmed Mpox case fatality rate in DRC, 1970-2024 *(based on the period before and after the Mpox global outbreak)*

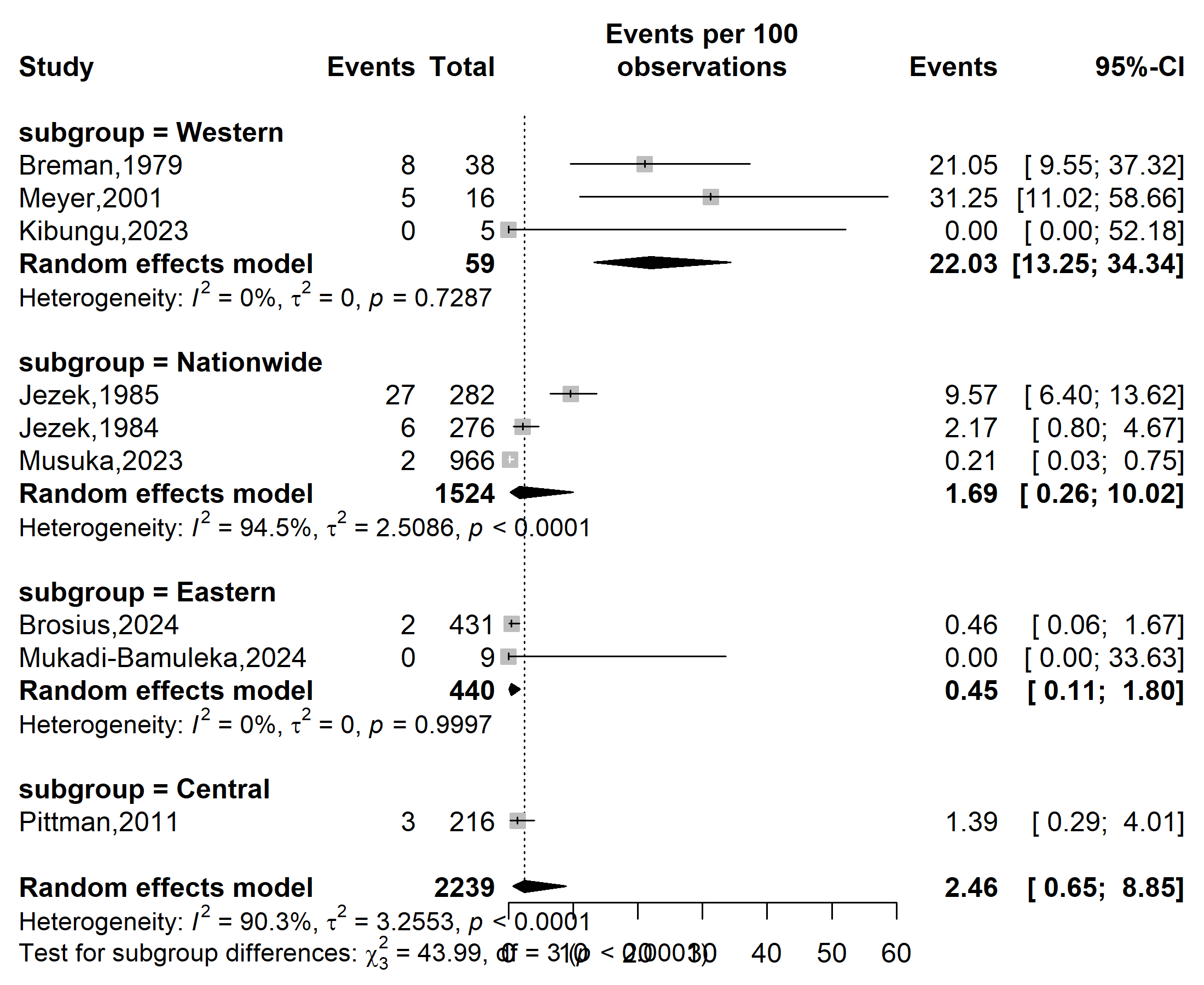

**Case fatality rate (%)**

**Event rate (%)**

Supplementary Fig. 2 Subgroup estimates of the confirmed Mpox case fatality rate in DRC, 1970-2024 *(based on geographical localization: Eastern: North Kiyu and South Kiyu; Central: Kasai Oriental and Sankuru; Western: Equateur and Kwango)*

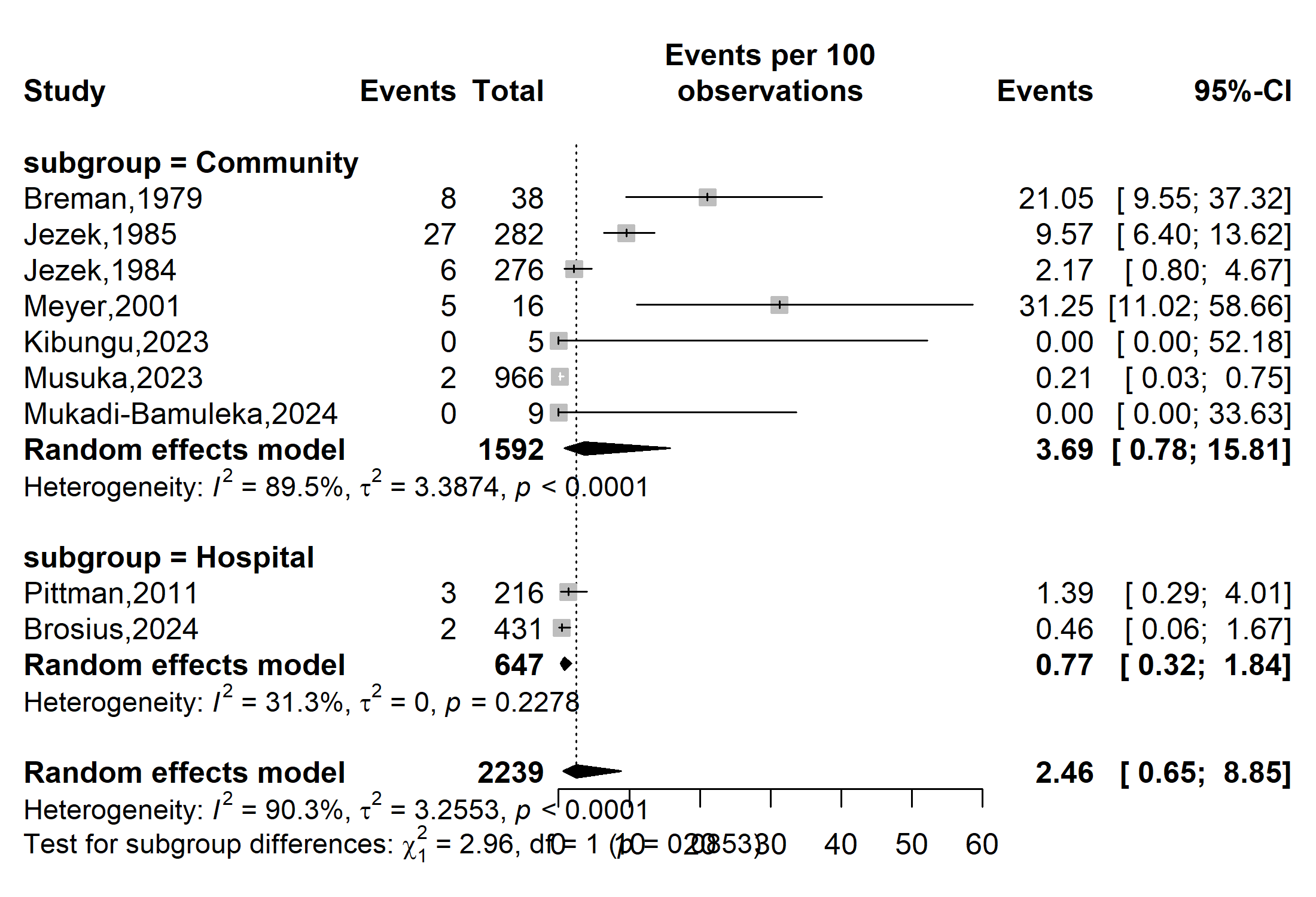

**Event rate (%)**

**Case fatality rate (%)**

Supplementary Fig. 3 Subgroup estimates of the confirmed Mpox case fatality rate in DRC, 1970-2024 *(based on study setting)*

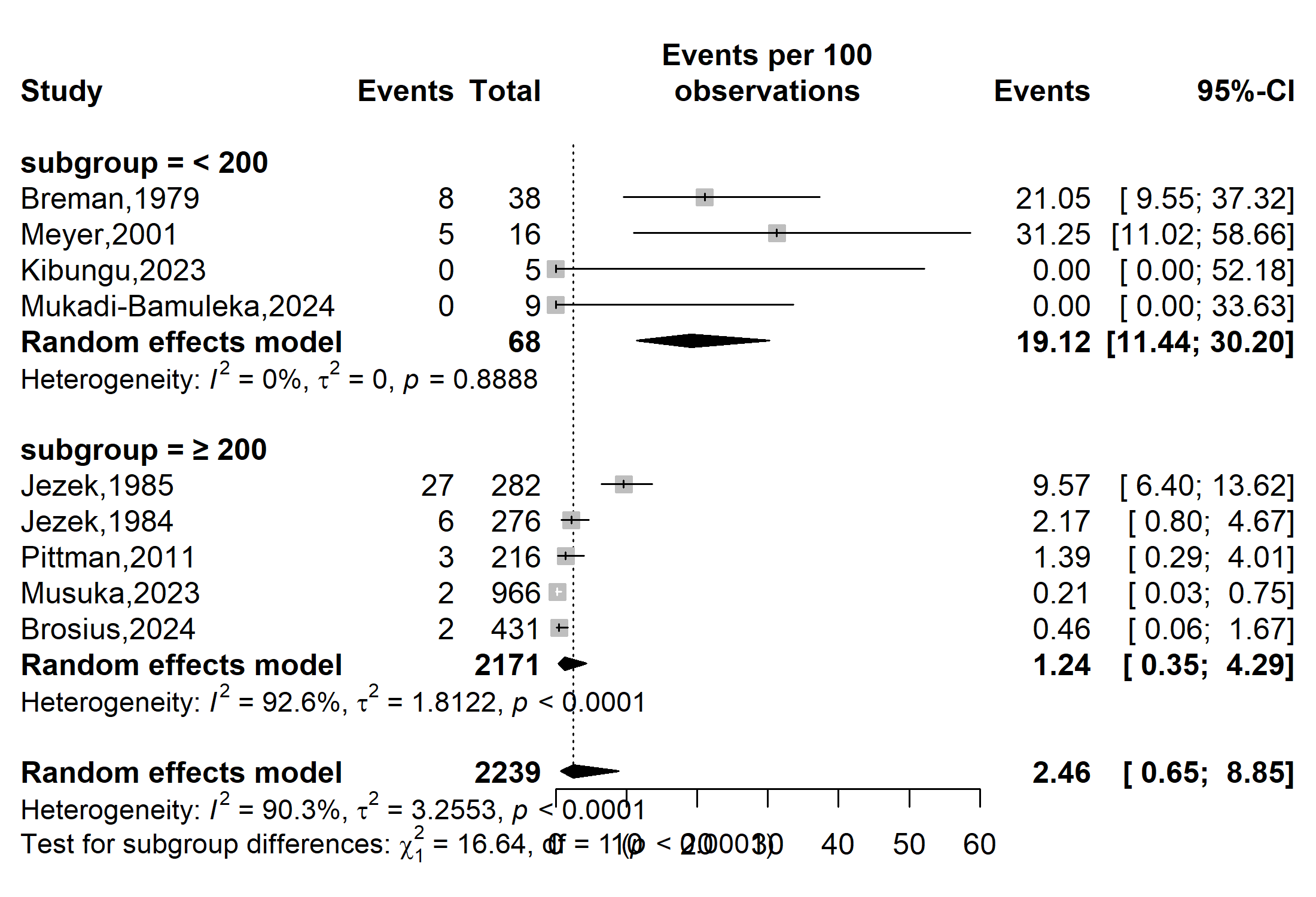

**Case fatality rate (%)**

**Event rate (%)**

Supplementary Fig. 4 Subgroup estimates of the confirmed Mpox case fatality rate in DRC, 1970-2024 *(based on the number of confirmed cases=disease burden)*

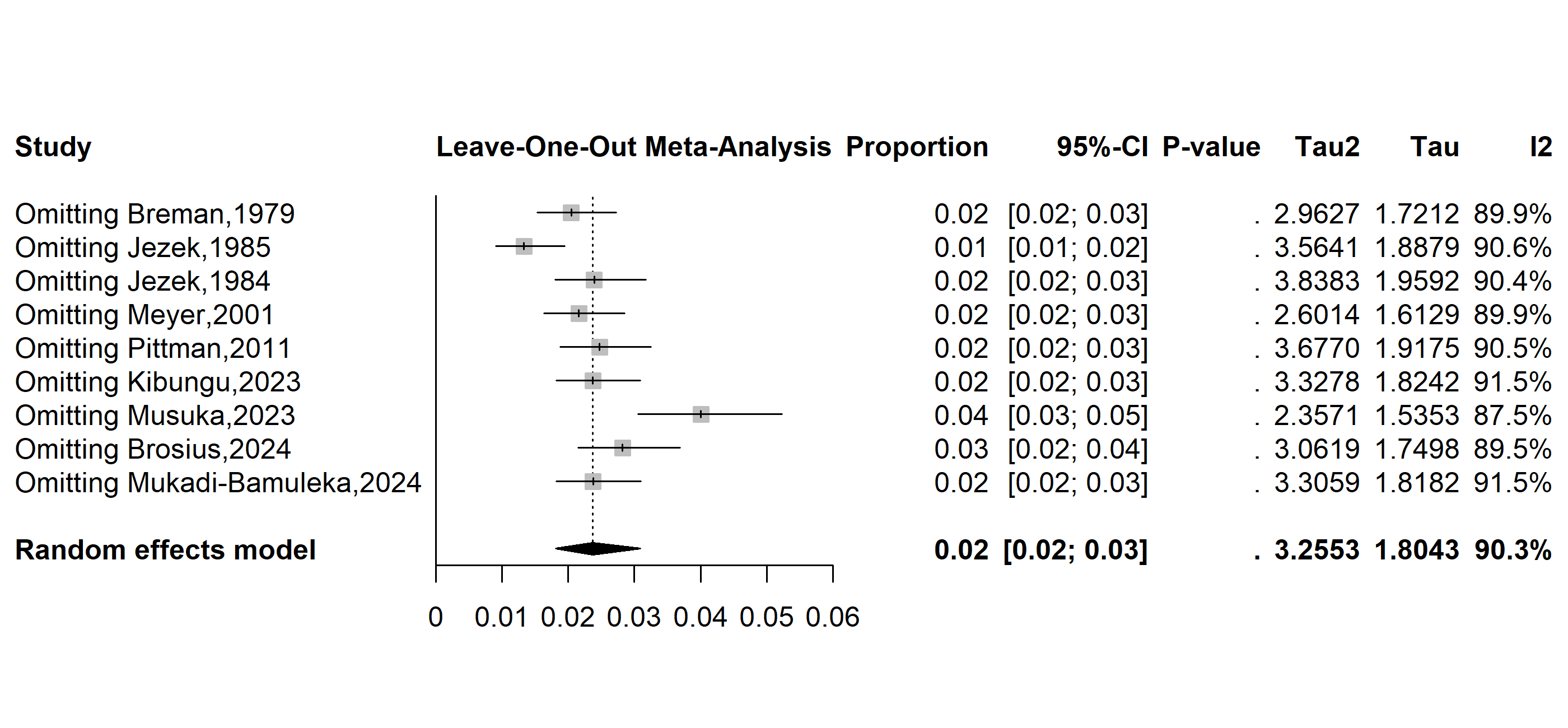

**Case fatality rate (%)**

**Event rate (%)**

Supplementary Fig. 5 Sensitivity analysis of the pooled case fatality rate estimates for confirmed Mpox cases in the DRC, 1970-2024

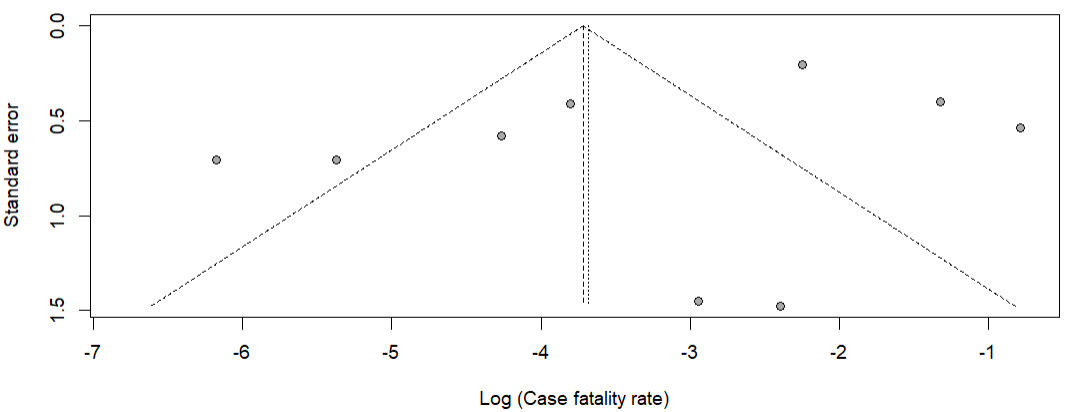

Supplementary Fig. 6 Funnel plot with pseudo 95% confidence limits and tests assessing the publication bias included studies
